# Evaluation of the replicability of systematic reviews with meta-analyses of the effects of health interventions

**DOI:** 10.1101/2025.07.17.25331623

**Authors:** Daniel G. Hamilton, Joanne E. McKenzie, Phi-Yen Nguyen, Melissa L. Rethlefsen, Steve McDonald, Sue E. Brennan, Fiona M. Fidler, Julian P. T. Higgins, Raju Kanukula, Sathya Karunananthan, Lara J. Maxwell, David Moher, Shinichi Nakagawa, David Nunan, Peter Tugwell, Vivian A. Welch, Matthew J Page

## Abstract

**Background:** Systematic reviews are often characterised as being inherently replicable but several studies have challenged this claim.

**Objectives:** To investigate the variation in results following independent replication of literature searches and meta-analyses of systematic reviews.

**Methods:** We included ten systematic reviews of the effects of health interventions published in November 2020. Two information specialists repeated the original database search strategies. Two experienced review authors screened full-text articles, extracted data, and calculated the results for the first reported meta-analysis. All replicators were initially blinded to the results of the original review. A meta-analysis was considered not ‘fully replicable’ if the original and replicated summary estimate or confidence interval width differed by more than 10%, and meaningfully different if there was a difference in the direction or statistical significance.

**Results:** The difference between the number of records retrieved by the original reviewers and the information specialists exceeded 10% in 25/43 (58%) searches for the first replicator and 21/43 (49%) searches for the second. Eight meta-analyses (80%, 95% CI: 49-96%) were initially classified as not fully replicable. After screening and data discrepancies were addressed, the number of meta-analyses classified as not fully replicable decreased to five (50%, 95% CI: 24-76%). Differences were classified as meaningful in one blinded replication (10%, 95% CI: 1-40%) and none of the unblinded replications (0%, 95% CI: 0-28%).

**Conclusions:** The results of systematic review processes were not always consistent when their reported methods were repeated. However, these inconsistencies seldom affected summary estimates from meta-analyses in a meaningful way.

**HIGHLIGHTS:** *What is already known on this topic:*

- Systematic reviews are often characterised as being inherently replicable, however, several studies have challenged this claim.
- Few studies have examined where and why inconsistencies arise, and what their impact is, when replicating multiple systematic review processes.

*What this study adds:*

- Replication of published systematic review processes (database searches, full-text screening, data extraction and meta-analysis) frequently produced results that were inconsistent with the original review.
- Following correction of replicator errors, the main drivers of variation in the results were incomplete reporting (e.g., unclear search methods, study eligibility criteria and methods for selecting study results) and reviewer data extraction errors.
- However, differences between the original reviewer’s and replicators’ summary estimates and confidence intervals were seldom meaningful.

## INTRODUCTION

The findings of systematic reviews have the potential to shape clinical practice and future research efforts in health and medicine in a profound way. However, despite their potential, there is growing unease among researchers about the reliability of systematic reviews and other published research [1–5]. This crisis of confidence is commonly referred to as the ‘replication crisis,’ or the ‘reproducibility crisis,’ and is often attributed to low replication success rates in large-scale replications of high-profile experiments across multiple scientific fields [6–10], including medicine [11–15].

Systematic reviews with meta-analyses are often characterised as being inherently replicable [16,17]. However, some evidence has brought this claim into question. For example, Rethlefsen et al. [18] found that when repeating 88 database searches from 39 systematic reviews, only 41 (47%) searches retrieved a similar number of records (i.e., differed by less than 10% to the original). Similarly discordant results have been observed when replicating other important review processes, such as article screening [19,20], data extraction [21–25], and risk of bias assessments [26,27]. Many of these studies also evaluated the impact of discrepancies in these processes on the findings of reported meta-analyses [19,21–25].

A limited number of studies have attempted to replicate multiple review processes for systematic reviews of health interventions [28,29]. The most relevant study [29] replicated multiple processes from eight systematic reviews (published between 1998 and 2007) that examined the effects of pharmacological interventions for irritable bowel syndrome. The authors found six reviews had missed 22 eligible randomized trials and incorrectly included 35 trials. They also identified 80 data extraction errors across all reviews. These discrepancies and errors resulted in at least a 10% change to the summary estimate in five of sixteen meta-analyses (31%), and a reversal of the statistical significance in four (25%).

Investigation of the replicability of multiple systematic review processes is important for determining where and why inconsistencies arise, and what their impact is. Given the limited number of replication studies that have been performed, we aimed to evaluate both the extent and source of variation in results when teams of information specialists and systematic reviewers independently replicated the database searches and meta-analyses of systematic reviews assessing the effects of health interventions.

## METHODS

This study was the fourth in a series of studies comprising the REPRISE (REProducibility and Replicability In Syntheses of Evidence) project [30–34]. An abbreviated version of the methods is provided here. The complete methods, including the list of deviations from the original study protocol [30] (Supplementary Table 1), are reported in the supplementary materials.

### Sampling frame

Systematic reviews selected for this study were sampled from those identified in the first study of the REPRISE project, which assessed the reporting completeness of 300 systematic reviews with meta-analyses published in November 2020 [32]. For this study, we selected for replication the reviews from the first REPRISE study which met the following inclusion criteria: 1) reported the full Boolean search strategy for each database searched; 2) reported the number of records retrieved by each search, 3) provided information on which studies were included in the review and in the first reported (i.e., “index”) pairwise meta-analysis; 4) reported summary statistics (e.g., means and standard deviations) or an effect estimate and associated measure of precision (e.g., standardised mean difference and 95% confidence interval) for each study in the index meta-analysis; and 5) did not include more than 10 studies in the index meta-analysis. For the current study, we also excluded reviews which had been retracted and restricted our attention to meta-analyses undertaken in the frequentist framework.

### Replication personnel

Two replications were completed in this study: 1) replications of the database literature searches and 2) replications of the index meta-analysis (including the full-text screening, data extraction and calculation steps). We did not replicate other review processes (e.g., title and abstract screening, risk of bias assessments), but used the results of the original reviewers when required. For example, when reviewers excluded studies based on risk of bias, we used the reviewers’ risk of bias assessment to determine study eligibility. A flow diagram depicting the procedure followed for each replication is shown in Figure 1. One author (MJP) with expertise in performing and evaluating the conduct and reporting of systematic reviews was responsible for the coordination of all replication activities. Replications of the literature searches were performed by two information specialists (MLR and SM), each with over 25 years’ experience developing search strategies for systematic reviews, clinical guidelines, and other evidence syntheses. Replication of the meta-analyses were performed by two researchers with experience screening articles, collecting and preparing outcome data, and performing meta-analyses for more than three systematic reviews within the past three years (DGH and P-YN). None of the four replicators were involved in the conduct of any of the ten systematic reviews selected for replication, any of the studies included within them, nor had performed a systematic review that addressed the same or similar questions. All replicators were instructed not to seek out any information on the ten reviews (e.g., a PROSPERO entry, a review protocol, the review report) prior to submission of the results of their initial replications (i.e., were blinded to the results of the reviews). The blinded replications were conducted to provide a test of the replicability of the methods as documented in the review report. We then followed this up with unblinded replications which allowed us to test the replicability of both the methods and results, as well as investigate the causes of observed discrepancies, including assessing the impact of errors introduced by the replication team.

**Figure 1.**
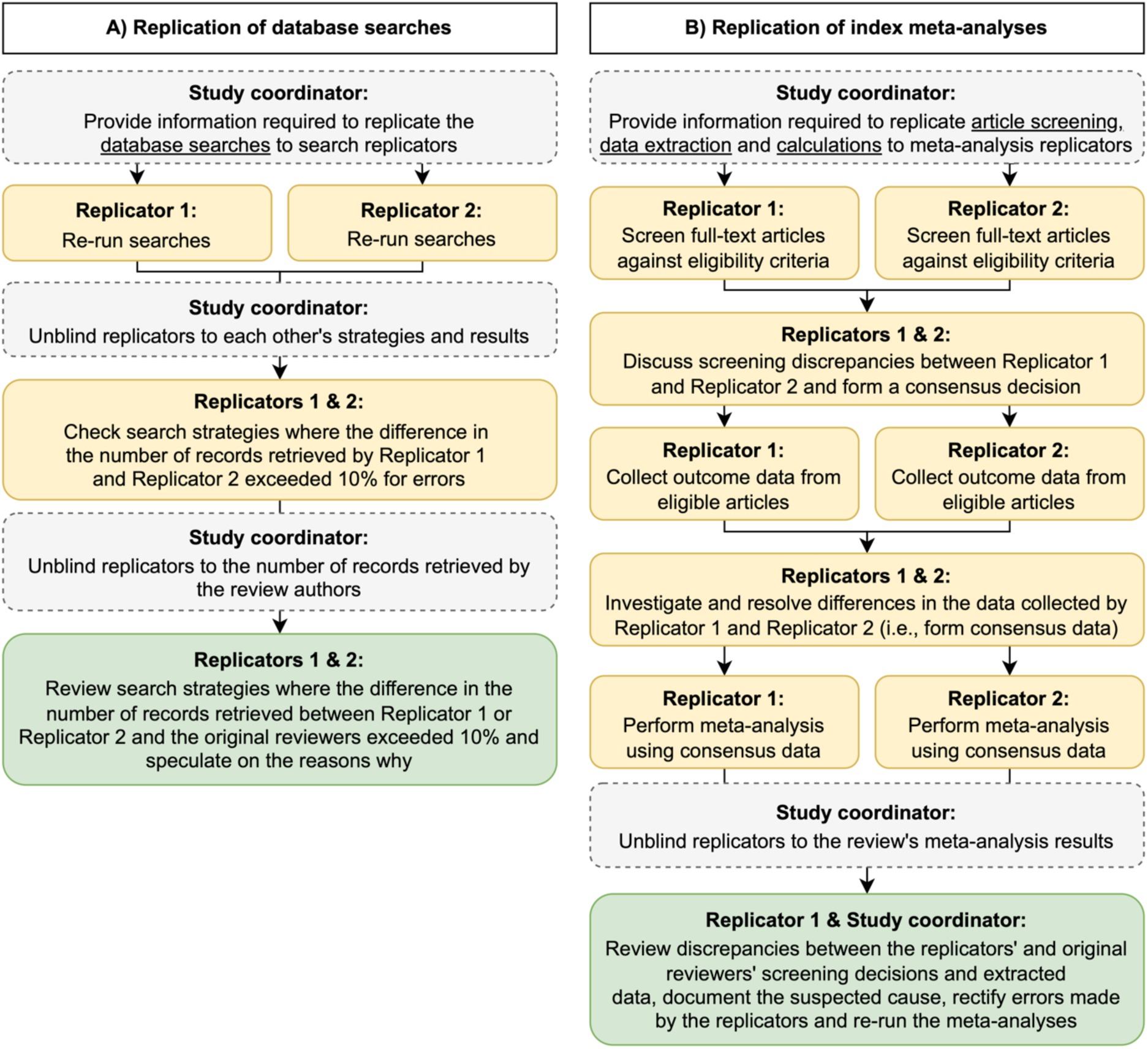
Flow diagram outlining the process followed to replicate the selected review’s (A) database searches and (B) index meta-analysis. Note that the yellow boxes refer to activities performed by replicators while blinded, and green boxes refer to activities occurring after unblinding.

### Extracted information for replicators

The study coordinator (MJP) assembled all information required to perform and evaluate the findings of the replications, which was subsequently checked for accuracy by one of the replicators (DGH) after they were unblinded. Exactly what information was provided to the replicators in the blinded phase and unblinded phase is reported in Table 1 and the project’s OSF page [35].

**Table 1.** Information extracted from selected systematic review reports and associated supplementary materials to guide replicators and compare results.

| Review process | Data item extracted | Data given to blinded replicators | Data given to unblinded replicators |
| --- | --- | --- | --- |
| Database searches | Line-by-line search strategy for each database consulted | Yes | Yes |
|  | The platform used | Yes | Yes |
|  | Date ranges when databases were last searched | Yes | Yes |
|  | The number of records yielded by each search | No | Yes |
| Full-text article screening | The question addressed by the index meta-analysis | Yes | Yes |
|  | Eligible participants, interventions, comparators, outcomes, study designs, languages of publication, report types for the index meta-analysis | Yes | Yes |
|  | Reports of studies that were included in the index meta-analysis | Yes | Yes |
|  | Reports of studies which were not included in the index meta-analysis (i.e., studies that were included in other analyses or excluded by the reviewers after full-text screening) | Yes | Yes |
|  | Original reviewers' screening decisions for each study | No | Yes |
| Data extraction | Decision rules reported by the original systematic reviewers regarding which results to select from studies (e.g., which measurement scale, time point, or analysis sample was selected) | Yes | Yes |
|  | Results of each study included in the index meta-analysis (i.e. summary statistics and effect estimate with a measure of precision) | No | Yes |
| Meta-analysis | The effect measure(s) used (e.g., risk ratio) | Yes | Yes |
|  | Meta-analysis method used (e.g., inverse-variance) | Yes | Yes |
|  | Meta-analysis model used (e.g., random effects) | Yes | Yes |
|  | Between-study variance estimator used (e.g., DerSimonian-Laird) | Yes | Yes |
|  | Analysis software used (e.g., RevMan) | Yes | Yes |
| | Results of the meta-analysis (i.e., summary estimate and measure of precision) and associated heterogeneity statistics (e.g., $\text{Chi}^2$ value, $I^2$, $\tau^2$) | No | Yes |

### Replication methods

We recognise that the terminology for ‘replication’ is not standardised within and across disciplines [36,37]. In this study, we adopted the non-procedural definitions of replication advocated by Nosek & Errington [38] and Machery [39]. That is, replicators did not need to follow every single step exactly as reported in the original systematic review, but they were constrained by the original review question and were instructed to avoid making changes to the methods and concepts that might be reasonably judged to violate an attempt to answer that question.

### Database search replications

For the literature search replications, replicators were asked to re-run all database search strategies using the same date limits as reported in the original review. When able, searches were run using the same platform and exact same search string and date limits as reported in the original review. Where possible, searches were also run so as not to retrieve records that were published within the relevant date range but indexed in the database after the original search was run. The two replicators performed all literature searches independently and documented which original search strategies they attempted to re-run, any assumptions made, and the number of records that each search yielded. Following submission of their search results, the replicators were unblinded to each other’s search strategies and results, then given the opportunity to amend their searches if any errors were detected. Next, the replicators were unblinded to the number of records retrieved by the original reviewers, and where the replicators’ search results differed from the original reviewers’ by more than 10%, were asked to speculate on the likely causes for the discrepancy.

### Index meta-analysis replications

For the meta-analysis replications, replicators were given the full-text reports of all articles included in the review (not just the index meta-analysis), as well as those cited as being excluded from the review, then asked to screen all reports against the eligibility criteria for the index meta-analysis. Replicators screened each article independently, recording their decision (‘include’, ‘exclude’, or ‘unsure’) and rationale, then met afterwards to discuss and resolve any discrepancies or uncertainties and form a consensus decision. In some situations, further information concerning the review’s eligibility criteria was requested from the study coordinator to aid decision-making (e.g., under what cut-off were doses considered “low dose” for Vitamin D supplementation), which was investigated and provided to the replicators when available in the systematic review report.

The data required for the meta-analysis were then independently extracted by both replicators from the body text, tables or plots in the study reports, or supplementary materials of the studies deemed eligible. WebPlotDigitizer (v4) was used to extract data from plots. The algorithm proposed by Guyot et al. (2012) [40] was executed in R to derive approximations of the hazard ratios and their standard error from data extracted from Kaplan-Meier curves. If the required data were not reported, one author (DGH) attempted to contact the primary study authors (i.e., authors of the study included in the systematic review) to obtain the required information. If the replicators were unable to source required data for an eligible study (including from the study authors), it was classified as having missing results.

Any discrepancies between numerical data extracted from text and tables in the study report by the replicators were discussed and resolved. Whereas, based on prior calibration testing, differences between replicators greater than 0.5 units for data extracted from plots where the length of the axis of interest ranged between 25 to 100 units, were investigated and investigated for possible errors (e.g., axis alignment errors, timepoint selection errors). Additionally, percentage differences between hazard ratios, upper and lower confidence interval bounds, and standard errors derived from Kaplan-Meier curves greater than 5% were investigated. Once all data derived from plots were finalised (including the estimation of hazard ratios), the arithmetic mean of the two values were calculated and used for the meta-analysis.

After consensus was reached on the extracted data, replicators independently performed the calculations to generate meta-analysis results. Given we did not have access to all software used by the reviewers, the replicators were instructed to use software they were most familiar with, namely, R (v4.3.1) using the meta package (v7.0.0) (DGH) and Stata 17 using the in-built meta command (P-YN). The results from both replicators are reported.

Following submission of the results of the article screening, data extraction, and calculation steps, the replicators were then unblinded, and discrepancies between the replicators and original reviewers in each step were identified and jointly investigated by two authors (DGH and MJP) to try to determine the cause. Where further data needed to be extracted (e.g., where an exclude decision was updated to include), this was performed by one author (DGH) and checked for accuracy by another (MJP). When the causes of discrepancies could not be confidently identified, one author (DGH) contacted the original reviewers to seek clarification. Once all discrepancies were reviewed and addressed, the results of the replications of each step were updated. These updated findings are referred to as the ‘unblinded’ results in the results section.

### Outcomes

The study’s outcomes are reported in Table 2. For the literature searches, we calculated the number of times the percentage difference between the number of records retrieved by the original reviewers and replicators was less than or equal to 10%. The cutoff of 10% was chosen to allow for expected variation in the number of records retrieved due to database changes, as well as comparability with previous related research [18]. Similarly, meta-analyses were classified as “fully replicable” if both the percentage difference between the original reviewers’ and replicators’ summary estimates *and* confidence interval widths did not exceed 10%. Furthermore, we classified replicated summary estimates as “meaningfully” different if they were opposite in direction to the original, or had a difference in the statistical significance, or both, given in practice, these differences might lead reviewers to reach different conclusions.

**Table 2.**
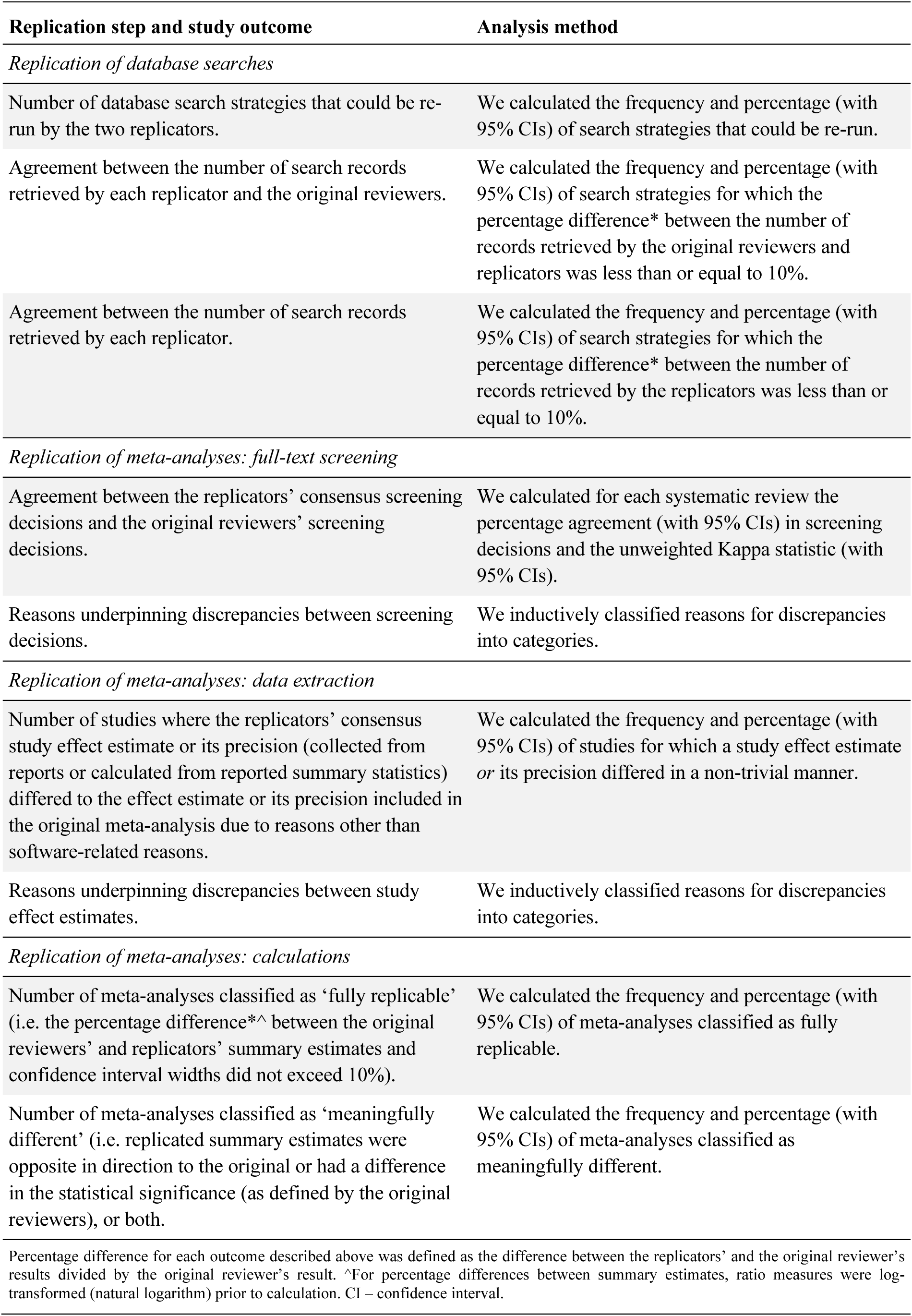
Study outcomes and analysis methods.

| Replication step and study outcome | Analysis method |
| --- | --- |
| *Replication of database searches* |  |
| Number of database search strategies that could be re-run by the two replicators. | We calculated the frequency and percentage (with 95% CIs) of search strategies that could be re-run. |
| Agreement between the number of search records retrieved by each replicator and the original reviewers. | We calculated the frequency and percentage (with 95% CIs) of search strategies for which the percentage difference* between the number of records retrieved by the original reviewers and replicators was less than or equal to 10%. |
| Agreement between the number of search records retrieved by each replicator. | We calculated the frequency and percentage (with 95% CIs) of search strategies for which the percentage difference* between the number of records retrieved by the replicators was less than or equal to 10%. |
| *Replication of meta-analyses: full-text screening* |  |
| Agreement between the replicators' consensus screening decisions and the original reviewers' screening decisions. | We calculated for each systematic review the percentage agreement (with 95% CIs) in screening decisions and the unweighted Kappa statistic (with 95% CIs). |
| Reasons underpinning discrepancies between screening decisions. | We inductively classified reasons for discrepancies into categories. |
| *Replication of meta-analyses: data extraction* |  |
| Number of studies where the replicators' consensus study effect estimate or its precision (collected from reports or calculated from reported summary statistics) differed to the effect estimate or its precision included in the original meta-analysis due to reasons other than software-related reasons. | We calculated the frequency and percentage (with 95% CIs) of studies for which a study effect estimate or its precision differed in a non-trivial manner. |
| Reasons underpinning discrepancies between study effect estimates. | We inductively classified reasons for discrepancies into categories. |
| *Replication of meta-analyses: calculations* |  |
| Number of meta-analyses classified as 'fully replicable' (i.e. the percentage difference*^ between the original reviewers' and replicators' summary estimates and confidence interval widths did not exceed 10%). | We calculated the frequency and percentage (with 95% CIs) of meta-analyses classified as fully replicable. |
| Number of meta-analyses classified as 'meaningfully different' (i.e. replicated summary estimates were opposite in direction to the original or had a difference in the statistical significance (as defined by the original reviewers), or both). | We calculated the frequency and percentage (with 95% CIs) of meta-analyses classified as meaningfully different. |
| Percentage difference for each outcome described above was defined as the difference between the replicators' and the original reviewer's results divided by the original reviewer's result. ^For percentage differences between summary estimates, ratio measures were log-transformed (natural logarithm) prior to calculation. CI – confidence interval. |  |

Percentage difference for each outcome described above was defined as the difference between the replicator’s and the original reviewer’s results divided by the original reviewer’s result. For percentage differences between summary estimates, ratio measures were log-transformed (natural logarithm) prior to calculation.

### Statistical analysis

Categorical data and continuous data are presented descriptively using i) counts and proportions, and ii) medians, interquartile ranges (IQR) and ranges, respectively. All analyses were performed in R (v4.3.1). Confidence intervals around proportions for binary outcomes were calculated using the modified Wilson interval method as proposed by Brown et al. [41] using the DescTools package (v0.99.55). Unweighted Kappa statistics and 95% confidence intervals were calculated using the psych package (v2.4.6.26). Figures were generated via draw.io (Figure 1), R using the ggplot2 (v3.5.1) package (Figure 2) and Microsoft Excel (Figure 3A & 3B). All data, code, and materials needed to reproduce the findings of this research have been made publicly available on the Open Science Framework (https://osf.io/v3mnh/) [35].

**Figure 2.**
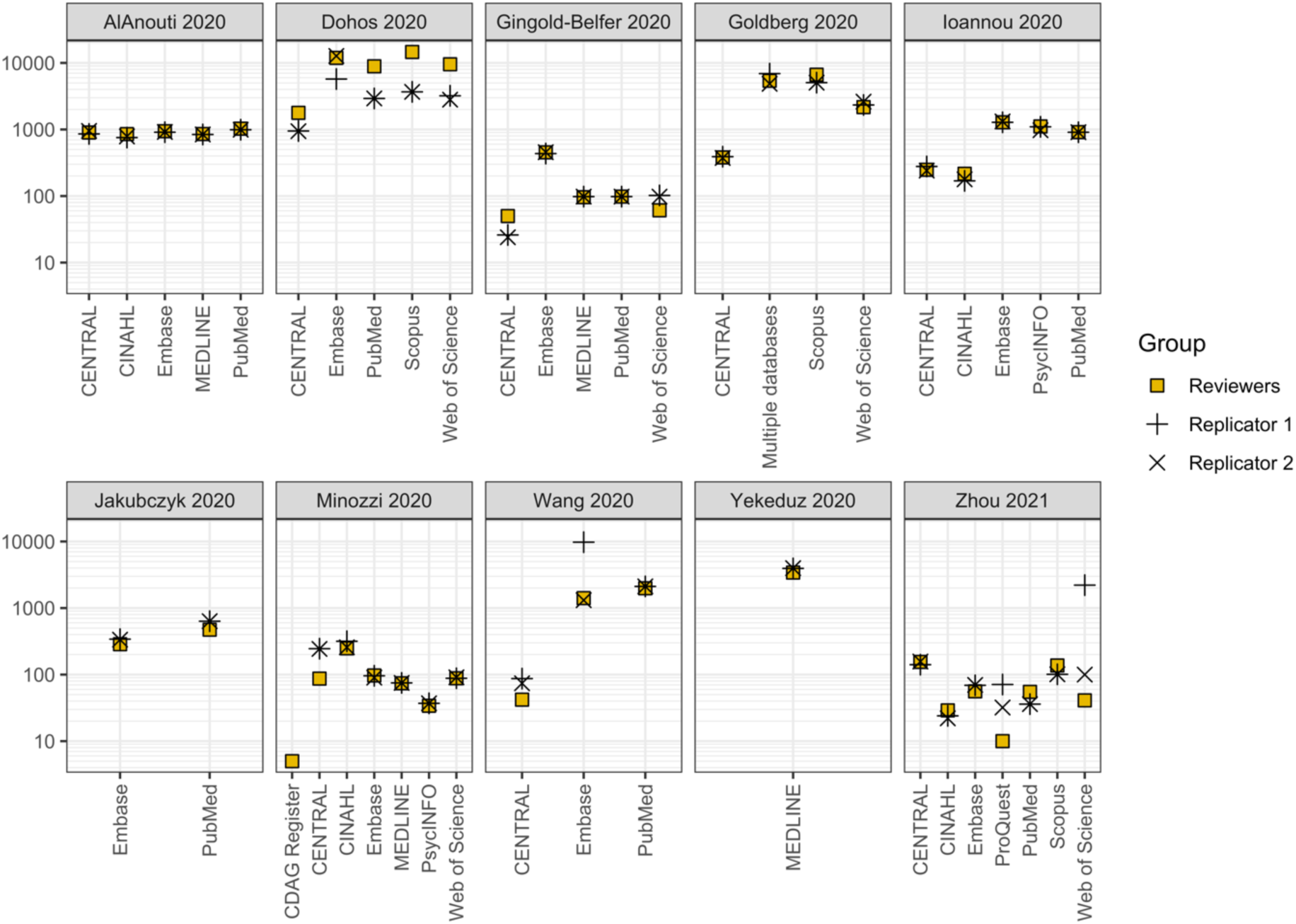
Scatter plot of the number of records retrieved by the original reviewers (square symbols) and the first (plus signs) and second (cross signs) replicators by systematic review. Note: These numbers for Replicators 1 and 2 refer to the records retrieved following correction of initial errors they had made. The y-axis is presented on a logarithmic scale (base 10).

**Figure 3A.**
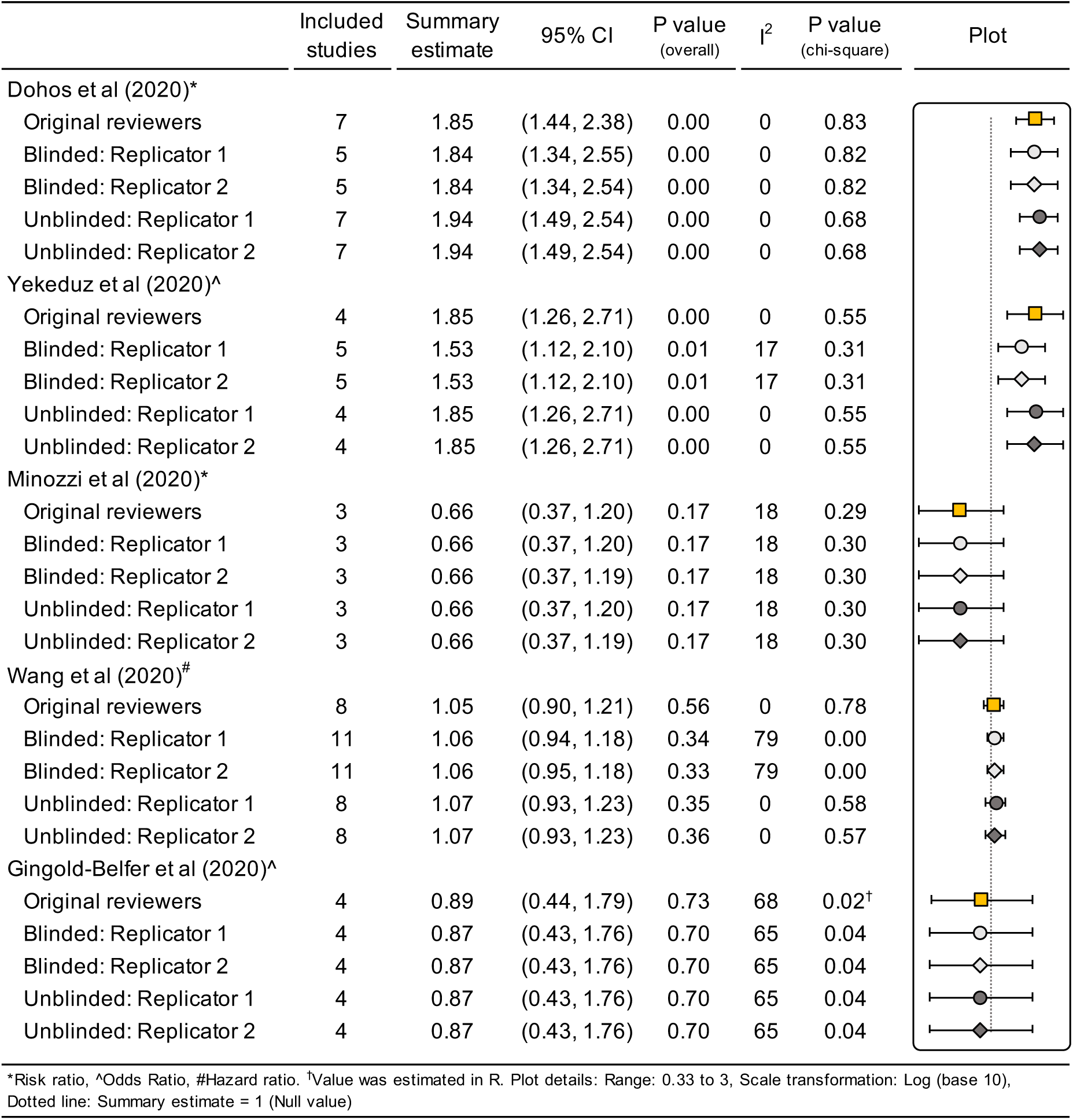
Replication of meta-analyses of ratio measures: Comparison of the results of the original reviewer’s and the replicators’ (blinded and unblinded) index meta-analyses.

**Figure 3B.**
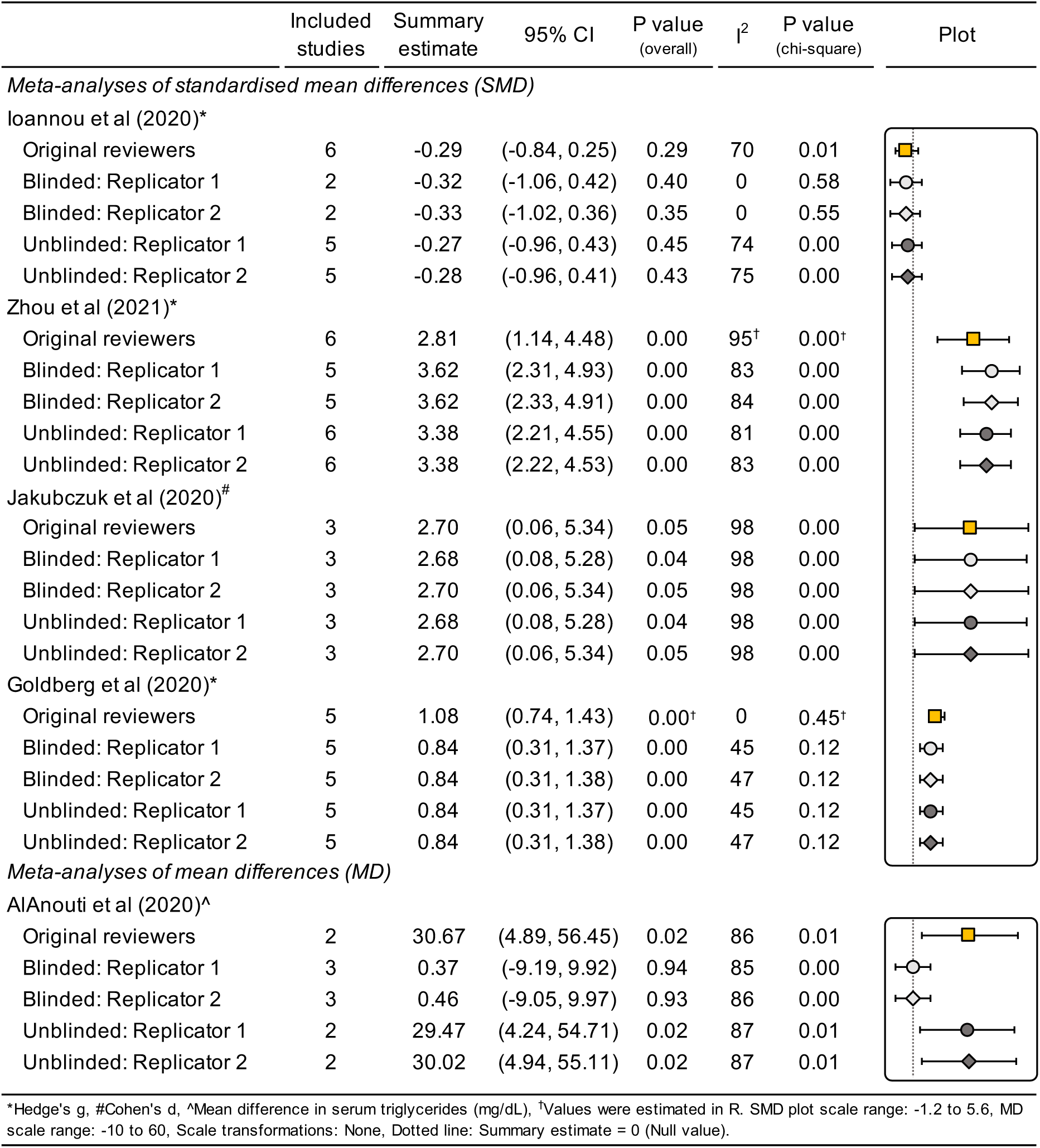
Replication of meta-analyses of difference measures: Comparison of the results of the original reviewer’s and the replicators’ (blinded and unblinded) index meta-analyses.

### Sensitivity analysis

We performed two post-hoc sensitivity analyses to investigate the robustness of two replicated meta-analyses. The first sensitivity analysis removed hazard ratios estimated using Guyot and colleague’s (2012) [40] algorithm from one meta-analysis that did not report the total number of events and the numbers at risk (other than at time zero) alongside the reported Kaplan-Meier curves. This was due to warnings provided the developers of the algorithm that its accuracy is likely to be poor (i.e., associated with a tenfold increase in the estimated mean absolute error) when the aforementioned information has not been provided. The second sensitivity analysis included a result from a study eligible for another meta-analysis where the effect estimate was calculated using means and standard deviations estimated from reported medians and ranges via the application of normal and non-normal data transformation methods [42,43] via the estmeansd package (v1.0.1) in R.

## RESULTS

### Characteristics of the included reviews

Ten of the 300 systematic reviews from the first REPRISE study met the inclusion criteria for this study [44–53]. The 10 reviews examined different interventions, including pharmaceuticals (e.g., methadone), behavioural interventions (e.g., sleep deprivation), surgical techniques (e.g., lymphadenectomy) and devices (e.g., laser therapy) (Table 3). Reviewers frequently limited eligible study designs to randomized trials (N=7/10, 70%). Half registered the review’s details or disseminated a protocol outlining the planned methods (N=5/10, 50%). The software used most often to perform meta-analyses was RevMan (N=5/10, 50%).

**Table 3.**
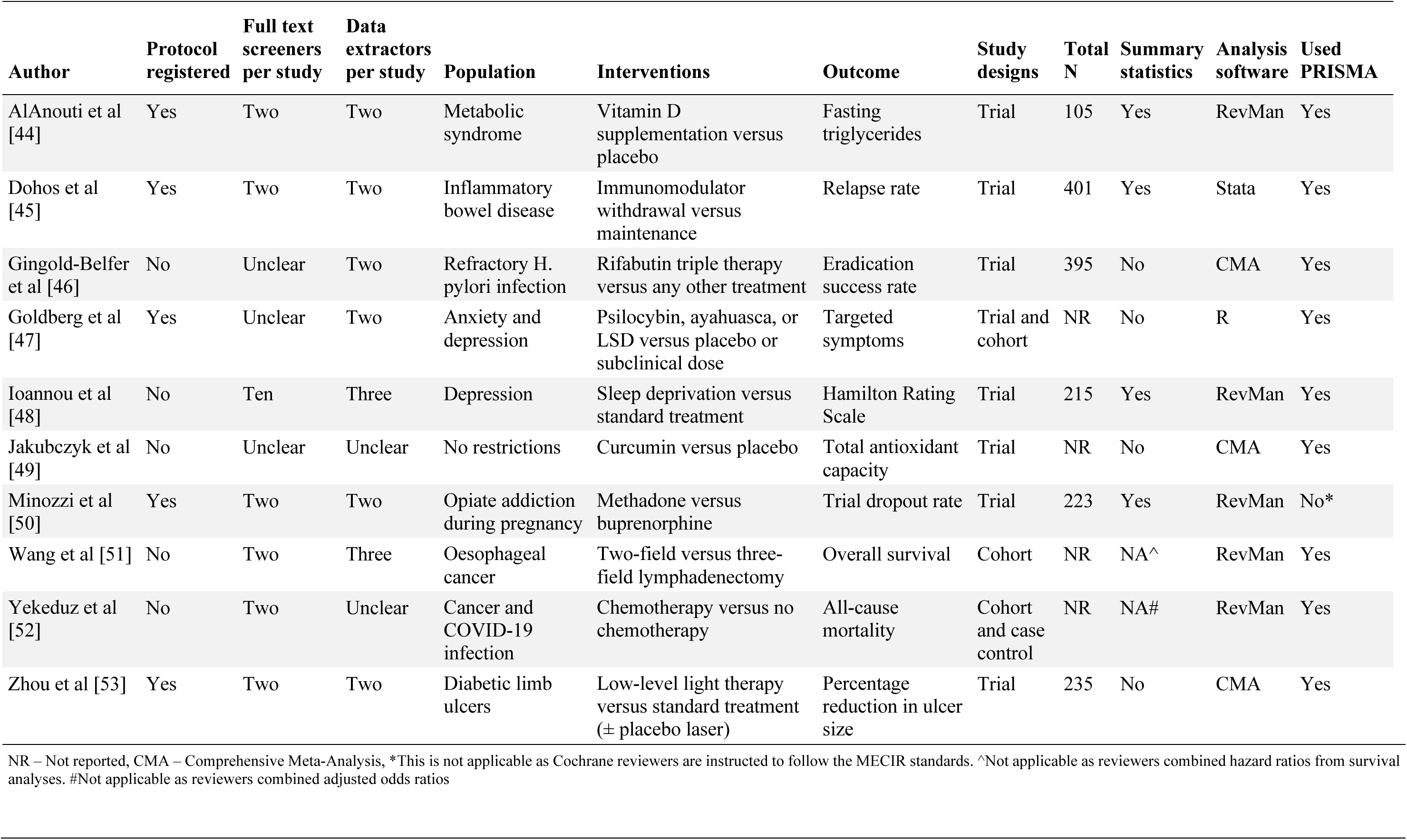
Characteristics of the reviews and index meta-analyses selected for replication.

### Replication of literature searches

Across the 10 reviews, 46 bibliographic database search strategies were reported in sufficient detail to enable replication. A median of five databases (IQR: 3.5 to 5.8, Range: 1 to 7) were searched per review, with the most frequently searched databases being CENTRAL, Embase and PubMed (all N=8/10, 80%). Both replicators were able to re-run all but one reported search strategy (N=45/46, 98%, 95% CI: 89-100%). The only search that could not be re-run was of the Cochrane Drugs and Alcohol Group’s Specialised Register as it was inaccessible to the public. The number of records retrieved by the original reviewers and replicators by review and database are shown in Figure 2 and Supplementary Figure 1, respectively.

For the 43 searches where the number of records retrieved by the original reviewers was reported, the percentage difference in number of records for the original reviewers and each of the two replicators exceeded 10% for approximately half of the searches (Search replicator 1: N=25/43, 58%, 95% CI: 43-72%; Search replicator 2: N=21/43, 49%, 95% CI: 35-63%). In contrast, the difference between the number of records retrieved by the two replicators exceeded 10% in 14 of 45 cases (31%, 95% CI: 20-46%). For no review or database did the results of all searches differ by less than 10%. The most common reasons for differences in the number of records retrieved between the replicators and original reviewers were thought to relate to syntax errors, undisclosed fields and limits, and differences in the platforms used, the database(s) searched within the platform and their default search settings (further details in Supplementary Table 2).

### Replication of meta-analyses: full text screening

In total, 185 articles were screened, of which 13 discrepancies occurred between the blinded replicators’ consensus decision and the original reviewers (κ=0.82, 95% CI: 0.72-0.91). At a review level, the replicators obtained perfect agreement or almost perfect agreement (i.e., a κ point estimate > 0.80) with the original reviewers in half the reviews (Table 4). Following unblinding, when the 13 screening discrepancies were investigated, they were deemed to be due to the study coordinator giving incomplete or incorrect eligibility criteria to the replicators (N=4/13, 31%), incomplete or ambiguous eligibility criteria in the review reports (N=4/13, 31%), errors by the replicators (N=2/13, 15%), replicators’ lack of content expertise (N=1/13, 8%), and unclear reasons (N=2/13, 15%). Overall, there were 10 instances where the replicators changed their screening decision after being unblinded (5 reclassified as eligible and 5 as ineligible), and three where they did not (1 eligible and 2 ineligible). Further details on the circumstances behind the differing screening decisions are outlined in Supplementary Table 3.

**Table 4.** Comparison of full-text article screening decisions between the reviewers and replicators’ consensus judgements.

| Author | Articles screened | Number of studies included in the index meta-analysis | | | Agreement between replicators' consensus judgements and original reviewers' judgements ($\kappa^*$, 95% CI) | | Percentage agreement between replicators' consensus judgements and original reviewers' judgements (% , 95% CI) | |
| --- | --- | --- | --- | --- | --- | --- | --- | --- |
|  |  | Original reviewers | Blinded replicators | Unblinded replicators | Blinded replicators | Unblinded replicators | Blinded replicators | Unblinded replicators |
| AlAnouti et al [44] | 7 | 2 | 3 | 2 | 0.70 (0.17, 1.00) | 1.00 (1.00, 1.00) | 86% (49%, 99%) | 100% (65%, 100%) |
| Dohos et al [45] | 19 | 7 | 5 | 7 | 0.76 (0.45, 1.00) | 1.00 (1.00, 1.00) | 89% (69%, 98%) | 100% (83%, 100%) |
| Gingold-Belfer et al [46] | 33 | 4 | 4 | 4 | 1.00 (1.00, 1.00) | 1.00 (1.00, 1.00) | 100% (90%, 100%) | 100% (90%, 100%) |
| Goldberg et al [47] | 24 | 5 | 5 | 5 | 1.00 (1.00, 1.00) | 1.00 (1.00, 1.00) | 100% (86%, 100%) | 100% (86%, 100%) |
| Ioannou et al [48] | 33 | 6 | 3 | 5 | 0.62 (0.24, 1.00) | 0.89 (0.68, 1.00) | 91% (76%, 97%) | 97% (85%, 100%) |
| Jakubczyk et al [49] | 4 | 3 | 3 | 3 | 1.00 (1.00, 1.00) | 1.00 (1.00, 1.00) | 100% (51%, 100%) | 100% (51%, 100%) |
| Minozzi et al [50] | 25 | 3 | 3 | 3 | 1.00 (1.00, 1.00) | 1.00 (1.00, 1.00) | 100% (87%, 100%) | 100% (87%, 100%) |
| Wang et al [51] | 12 | 8 | 11 | 8 | 0.31 (-0.18, 0.80) | 1.00 (1.00, 1.00) | 75% (47%, 91%) | 100% (76%, 100%) |
| Yekeduz et al [52] | 16 | 4 | 5 | 4 | 0.85 (0.56, 1.00) | 1.00 (1.00, 1.00) | 94% (72%, 100%) | 100% (81%, 100%) |
| Zhou et al [53] | 12 | 6 | 5 | 6 | 0.50 (0.02, 0.98) | 0.67 (0.24, 1.00) | 75% (47%, 91%) | 83% (55%, 97%) |

\*Kappa coefficients ( $\kappa$ ) were interpreted as poor ( $\leq 0.00$ ), slight (0.01-0.20), fair (0.21-0.40), moderate (0.41-0.60), substantial (0.61-0.80), almost perfect (0.81-0.99) or perfect (1.00). CI – confidence interval.

### Replication of meta-analyses: data extraction

Data were extracted from the 46 reports identified as eligible during the blinded phase, and the six reclassified as eligible in the unblinded phase. Data were mostly extracted directly from study reports (N=35/52, 67%) or estimated from plots (N=14/52, 27%). For the 40 reports which had data extracted by both the original reviewers and the blinded replicators, 27 discrepancies were observed. The reasons underpinning the observed discrepancies were deemed to be due to trivial differences related to the analysis software used (N=9), variation when extracting data from plots (N=7), incomplete methods for selecting study results in the review report (N=7), reviewer errors (e.g., double counting of participants) (N=3), and an unknown reason (N=1). For the data extracted from studies reclassified as eligible in the unblinded phase, there were two discrepancies, one trivial software-related difference and one deemed to be due to under-specification of the methods for selecting study results in the review report. In total, 19 out of the 46 effect estimates and estimates of precision extracted by both the replicators (blinded and unblinded) and the original reviewers were classified as having differed due to non-software related reasons (41%, 95% CI: 28-56%). Further details on the circumstances behind these 19 discrepancies are outlined in Supplementary Table 4.

### Replication of meta-analyses: calculations

Both replicators were able to generate results for all 10 index meta-analyses using the data they extracted. Results of the original and replicated meta-analyses using data from the blinded and unblinded phases of the study are presented in Figures 3A and 3B (refer to Supplementary Figures 2 to 11 and Supplementary Table 5 for more detailed results for each review). Using the data collected during the phase of the study where the replicators were blinded to the reviews’ results, both replicators classified eight of 10 meta-analyses (80%, 95% CI: 49-96%) as not fully replicable (i.e., summary estimate or confidence interval length differed by more than 10% different to the original). One of ten meta-analyses (10%, 95% CI: 1-40%) was classified as meaningfully different from the original due to a difference in the statistical significance (Supplementary Table 6). This change in statistical significance was caused by the incorrect exclusion of a study by the replicators who were not informed by the study coordinator that the study was excluded due to the follow up duration being too much longer than the other included studies.

When the 10 meta-analyses were performed with the unblinded data (i.e., data after discrepancies between the original reviewers and replicators were identified and addressed, and only reviewer errors, or unresolvable or trivial software-related discrepancies remained), the number of meta-analyses classified as not fully replicable decreased from eight to five (50%, 95% CI: 24-76%) and the number considered meaningfully different from the original decreased from one to zero (0%, 95% CI: 0-28%). Numerous factors contributed to the ultimate variation observed between the original reviewers’ and blinded and unblinded replicators’ summary estimates (Supplementary Tables 7 and 8). After correction of replicator errors, the main drivers of variation in the summary estimates from the unblinded replications were unclear or ambiguous eligibility criteria in the review report, under-specification of the methods for selecting study results, and reviewer data extraction errors.

For the sensitivity analyses that removed potentially unreliable hazard ratios from one meta-analysis and included estimated summary statistics for another (Supplementary Figures 12 and 13), the number of meta-analyses based on the blinded data that were classified as not fully replicable did not change. However, summary estimates from the unblinded replications were not robust to the sensitivity analyses, with the percentage classified as not fully replicable increasing from 50% to 60%. The percentage of replicated meta-analyses considered to be meaningfully different from the original also increased from 0% to 20% due to the first summary estimate changing from statistically significant to non-significant and the second reversing direction.

## DISCUSSION

The current study found variation in results when replicating the literature searches and first reported meta-analyses of 10 systematic reviews, which is consistent with previous research. For the literature search replications, while all but one could be successfully re-run, the replicators were only able to retrieve a similar number of records for around half the searches (49% and 58%), which is consistent with two other similar studies (47% and 33%, respectively) [18,28]. The percentage of discrepancies occurring in the data extraction step that were determined to be due to errors made by the reviewers (11%, N=3/27) is consistent with an investigation of data extraction errors within 201 systematic reviews of adverse events [25] (17%, N=1762/10386). Furthermore, when applying our definitions for replication success to the studies by Ford et al. [29], Carrol et al. [24] and Xu et al. [25], they would have classified 31%, 50% and 42% of their meta-analyses of binary data as not fully replicable, respectively (our estimate is 50%). The same studies would have also only classified 25%, 0% and less than 10% as ‘meaningfully’ different from the original meta-analysis, respectively (our estimate is 0%). Similar findings have also been observed among replications of meta-analyses of continuous and ordinal data [22,23]

### Implications of the research

When reflecting on the causes of variation observed when replicating the literature searches and meta-analyses of systematic reviews, and how it might be minimised, three key implications emerge from our research. Our findings suggest that most reported database searches are likely not replicable, and that replication success is likely to be largely driven by the reporting completeness of the search strategy. Therefore, it is likely that search replicability could be enhanced if key details outlined in the PRISMA 2020 [54] and PRISMA-S [55] statements (e.g., exact databases and platforms searched, field and limits used) were more routinely reported.

Inadequate specification of the eligibility criteria for each synthesis, along with guidance on how to select results when there is multiplicity, explained the variation between the original reviewers’ and replicators’ summary estimates in some cases. The influence of incomplete reporting of eligibility criteria on replicability has been previously foreshadowed [56,57]. For example, in a recent evaluation of 41 systematic reviews of health interventions by Cumpston et al. [57], the authors estimated that less than a third of the reviews provided enough information to allow someone to replicate decisions about which included studies were eligible for each synthesis. In our sample, we note that the provision of information that clarified acceptable and unacceptable co-interventions, outlined rules for selecting results, and avoided ambiguous/conflicting instructions would have led to greater consistency between the original reviewers’ and replicators’ results. Such issues could be addressed through greater uptake of resources which assist review authors with developing and reporting their research questions and eligibility criteria for all planned and performed syntheses, such as the Cochrane Handbook for Systematic Reviews of Interventions [58] and the recently developed InSynQ (Intervention Synthesis Questions) checklist [59].

Lastly, more thorough data documentation practices (e.g., archival of, at minimum, the extracted summary statistics) might have yielded more consistent outcomes between the replicators and original reviewers. For example, greater availability of summary statistics from the studies included in the meta-analyses would likely have helped the replicators resolve discrepancies between replicators and original reviewers, particularly those related to the selection of results. However, improvements to documentation practices likely require top-down intervention (e.g., journal, funder, or institutional mandates) given previous research has shown, in the absence of a mandatory sharing policy, less than 1% of systematic reviewers make their data publicly available [60].

### Strengths and limitations

The current study had several strengths. We improved on previous replication research [21,22,25,28,29] by initially blinding the replicators to ensure they were not influenced by the original reviewers’ results. Importantly, in contrast to replication work involving participants, where experimental set ups can be sensitive to seemingly innocuous contextual factors (i.e., ‘hidden moderators’) [61], we were able to directly observe, investigate and resolve discrepancies, often with the assistance of the original reviewers. However, we also note several limitations. Firstly, though consistent with the study by Ford and colleagues [29], for logistical reasons our sample size was small, which has led to large imprecision in our estimates of the percentage of meta-analysis results that were fully replicable or were meaningfully different to the original. Information initially provided to the replicators was collected by one investigator only which resulted in the introduction of some errors. However, these errors were identified and addressed through the unblinded analysis. The study adopted a crude measure to classify ‘meaningful’ differences in summary estimates from meta-analyses which ignores clinically important changes in effect size and precision that could impact the certainty of evidence. However, given the minor changes in the summary estimates and confidence intervals overall, it is unlikely that our conclusions would have changed had we used a measure which factored in minimally important differences. The study also restricted its scope to the first reported meta-analysis of systematic reviews that met a high threshold for reporting quality. Therefore, our results may not generalise to reviews that have less complete reporting. We also did not replicate other important review processes (e.g., title and abstract screening) and at times relied on information reported by the original reviewers to guide our decision-making (e.g., risk of bias judgements).

### Future directions

For this study, we adopted language consistent with most systematic reviewers’ understanding of the term ‘replication’ [34] as well as definitions proposed by the National Academies of Sciences, Engineering, and Medicine [62]. However, we note that the terms ‘replicability’ and ‘reproducibility’ are still often used interchangeably [34,63]. Consequently, as suggested by others [34,63,64], standardisation of these terms, in addition to the continued discussion of other important challenges associated with performing replication studies, such as conflicts of interest [34,65,66], shortage of funding opportunities [34,67,68], determining when replications are informative and when they are redundant [64,69,70], and addressing difficulties associated with publishing them [34,66] should also be a research priority. We also highlight a lack of evidence on the replicability of other evidence synthesis methods (both quantitative and qualitative) beyond pairwise meta-analysis of aggregate data. There is also a need to assess the replicability of AI-supported/generated reviews given their increasing prevalence in the literature [71].

Finally, whilst the inability to replicate published findings has been a focal point of the replication crisis, it is but only one factor that contributes to our confidence in the reliability of a scientific claim. For example, as Errington et al. [15] keenly note, “a finding can be both replicable and invalid at the same time”. Therefore, while we have shown that summary estimates from meta-analyses were seldom ‘meaningfully’ affected by discrepancies occurring in earlier stages of the review, we have not demonstrated that the summary estimates themselves are valid [72]. As such, the performance of replications that address methodological shortcomings observed in original reviews would provide another useful line of research into the reliability of evidence generated from systematic reviews.

## CONCLUSION

Our results show that the results of systematic review processes are not always consistent when their methods were repeated. However, these inconsistencies seldom affected summary estimates from meta-analyses in a meaningful way. Lack of detail about the search methods, eligibility criteria and methods concerning which results to extract from eligible studies were impediments to replicability. Future work should investigate methods to improve author engagement with resources designed to improve the reporting of systematic reviews.

## Data Availability

All data, code and materials associated with this project are publicly available on the Open Science Framework under a CC-BY license (DOI: 10.17605/OSF.IO/V3MNH).

https://www.doi.org/10.17605/OSF.IO/V3MNH

## DECLARATIONS

### Ethics approval and consent to participate

Ethics approval was not required for this research.

### CRediT statement

Conceptualization: JEM, MJP; Data curation: DGH, MJP; Formal Analysis: DGH, JEM, PN, MJP; Funding acquisition: MJP; Investigation: DGH, PN, MLR, SM, MJP; Methodology: DGH, JEM, PN, MLR, SM, FMF, JPH, RK, SK, LJM, DM, SN, DN, PT, VAW, MJP; Project administration: DGH, MJP; Resources: DGH, JEM, PN, MLR, SM, MJP; Software: DGH, PN; Supervision: JEM, MJP; Validation: DGH; Visualization: DGH, JEM, MJP; Writing – original draft: DGH; Writing – review & editing: DGH, JEM, PN, MLR, SM, FMF, JPH, RK, SK, LJM, DM, SN, DN, PT, VAW, MJP.

