## Supplementary Materials for "Evaluation of the replicability of systematic reviews with meta-analyses of the effects of health interventions"

### SUPPLEMENTARY TABLES

Supplementary Table 1. Deviations from the planned methodology.

| Section | Original methods | Revised methods | Reason for modification |
|---|---|---|---|
| Sampling frame | We will restrict the sampling frame to systematic reviews that included 5-10 studies in the index meta-analysis. | We restricted the sampling frame to systematic reviews that included 2-10 studies in the index meta-analysis. | Increasing the range of studies that could be included in the index meta-analyses gave us a larger sample to draw from (227 of the 300 systematic reviews in REPRISE Study 1 had between 2 and 10 studies in the index meta-analysis, whereas 130 systematic reviews had between 5 and 10 studies in the index meta-analysis). Furthermore, restricting eligibility to systematic reviews with 5-10 studies in the index meta-analysis would have limited the generalisability of the sample. |
| Sampling frame | We will include systematic reviews in the sampling frame regardless of how completely the search methods and results were reported. | We restricted the sampling frame to systematic reviews in which authors reported the full Boolean search strategy for each database searched and the number of results yielded by each database (or a combination of them). | We had originally planned for replicators to attempt to construct search strategies from incompletely reported information (e.g. list of keywords only). We subsequently decided that it was more valuable to determine whether completely reported search strategies could be replicated, because if not, chances of replication success for incompletely reported ones would likely be even lower. Furthermore, for systematic reviews in which authors did not report the number of records yielded per database, we would be unable to compare the number of records yielded originally and upon replication. |
| Sampling frame | We will draw a random sample of 32 systematic reviews for replication. | We planned to draw a random sample of 10 systematic reviews for replication. | Doing so was subsequently considered more feasible. Unexpectedly, only 10 systematic reviews met the |

| Section | Original methods | Revised methods | Reason for modification |
|---|---|---|---|
| | | | eligibility criteria for inclusion, so we did not need to draw a random sample. |
| Crowdsourcing of reviewers | We planned to recruit 60 replicators via various avenues, with each team including at least 2 members. Each team would conduct 2 replications each. | We recruited 2 information specialists and 2 systematic reviewers who had previously worked closely with the study coordinator to conduct all 10 replications. | Doing so gave the study coordinator confidence that the replicators had the necessary expertise to carry out all tasks. |
| Data collection | We will share all instructions and PDF copies of full-text reports with replicators via the Open Science Framework repository. | Instructions were shared via Google Drive instead. | For convenience, as all replicators routinely use Google Drive in their day-to-day work. |
| Data collection | One of the REPRISE investigators (the study coordinator) will assemble all the information and files necessary for the replication of the 32 systematic reviews. | After completing all replications blinded to the results of the original review ("blinded replications"), one of the replicators checked the information extracted by the study coordinator for accuracy. | Doing so ensured that replications conducted after results of the original review were known ("unblinded replications") were guided by accurate information. |
| Data collection | We will also invite the authors of the original systematic reviews to provide us with a file containing all their screening decisions, if not already made publicly accessible. | Authors of the original systematic reviews were not contacted to provide such information. | We planned to use this information from the original reviewers to compare their title/abstract screening decisions with the replicators' decisions. However, we subsequently made the decision to not attempt to replicate the title/abstract screening phase of the systematic reviews, due to the substantial time investment required to do so. |
| Replication methods | Of the 32 reviews included, 30 will be replicated by one team each (with detailed information provided about the methods). The remaining two reviews will each be replicated by 15 teams (with | We decided not to perform the Many Analysts replications. | Not doing the Many Analysts replications would save time and resources. |

| Section | Original methods | Revised methods | Reason for modification |
|---|---|---|---|
| | minimal information provided about the methods) in a Many-Analysts style. | | |
| Replication methods | If full search strategies are not reported, replicators will be permitted to request the strategies from the authors of the original systematic review (with the contact made via one of the REPRISE investigators). | Replicators of the search will not contact authors of the original systematic review for non-reported search strategies. | There was no need to contact the original reviewers for the non-reported search strategies, because systematic reviews were only included if all search strategies were fully reported. |
| Replication methods | Replicators will document…any errors detected when rerunning each original search strategy…. | Replicators were not prompted via the replicator form given to them to record any errors detected when rerunning each original search strategy. | This was accidentally omitted from the replicator form by the study coordinator. However, the search replicators did end up recording errors they encountered on the form. |
| Replication methods | Replicators will document…the number of unique citations after duplicates were removed. | Replicators will not document the number of unique citations after duplicates were removed. | To obtain such information would have required the search replicators to download the number of citations yielded by each database and then remove duplicates via software. Doing so would have taken a long time in many cases (e.g. for databases where citations can only be exported in batches of 1000, and the total number of citations yielded is 15000), so we removed this requirement to save the search replicators' time. |
| Replication methods | Two team members will screen independently a random sample of a maximum of 100 titles and abstracts yielded from the searches against the inclusion criteria reported in the original review, and record their screening decisions ('include', 'exclude', or 'unsure'). | No screening of titles and abstracts yielded from the searches occurred. | We subsequently decided that replicating the screening of 100 titles and abstracts was a waste of time. We assumed that in many cases, all the titles and abstracts screened would be ineligible, which would have prevented us from people able to use agreement statistics (e.g. Kappa) to determine replication success. |

| Section | Original methods | Revised methods | Reason for modification |
|---|---|---|---|
| Replication methods | 'Results fully replicable' was originally defined as "no difference [with allowance for trivial discrepancies such as those due to computational algorithms] is observed between the original and recalculated meta-analytic effect estimate, its 95% confidence interval and inferences about heterogeneity reported in the original review); | Meta-analyses where both the original and replicated summary estimate of the and its 95% confidence interval width differed by 10% or less were classified as "results fully replicable". | We subsequently decided to use a cutoff value to remove subjectivity in the judgements. |
| Replication methods | A REPRISE investigator and the team will also independently specify whether they believe the observed difference between the original and recalculated summary estimate and its precision was meaningful, that is, would lead to a change in the interpretation of the results (classified as 'difference meaningful' or 'difference not meaningful'). | A difference between the original and replicated summary estimate was considered "meaningful" if either the statistical significance or direction differed. | We subsequently decided to use objective criteria to remove subjectivity in the judgements. |
| Interview methods | Once teams complete both replications, investigators will conduct semi-structured interviews (of approximately 30 min duration) with replicators to discuss the analytical steps they took and to understand the decision-making processes used when synthesising the data. | No interviews were conducted. | The interviews were designed mainly to capture replicators thoughts about the findings of the Many Analysts replications. Given these replications were not done, the interviews were considered less of a priority. |
| Data analysis | We will assess agreement between the original and replicated review in the number of citations yielded from each database, in total and once duplicates were removed, by calculating the weighted Kappa statistic and percentage agreement (both metrics will be presented with 95% confidence intervals). | We determined agreement in search results by calculated the percentage difference between the original reviewer's and replicators' search results. Percentage differences less than or equal to 10% were considered a successful replication. | Our plan to calculate a weighted Kappa statistic for search results was written in error. We used a 10% cutoff for comparability with a similar study (PMID: 38052277). |

| Section | Original methods | Revised methods | Reason for modification |
|---|---|---|---|
| Data analysis | We will assess agreement between the original and replicated review in screening decisions (where available) for the subset of titles and abstracts and full-text reports screened by replicators by calculating the weighted Kappa statistic and percentage agreement (both metrics will be presented with 95% confidence intervals). | We calculated an unweighted Kappa statistic and percentage agreement to assess agreement between the original and replicated screening decision for full text reports. | Titles and abstracts were not screened by replicators. We calculated unweighted Kappa because there were only two response options for screening: "Include" or "Exclude". |
| Data analysis | We will calculate the frequency and percentage (with 95% confidence intervals) of …, (ii) search strategies replicators needed to reconstruct or adapt based on the partial information available | We did not calculate the frequency of search strategies replicators needed to reconstruct or adapt based on the partial information available. | None of the search strategies replicated needed to be reconstructed or adapted based on partial information available, as all were completely reported. |
| Data analysis | We will calculate agreement between the original and replicated meta-analytic effects, displayed using Bland-Altman plots. | We did not calculate agreement between the original and replicated summary estimates using Bland-Altman plots. | We subsequently considered Bland-Altman plots would no longer be useful to present given the smaller number of systematic reviews included (n=10) and the variation in effect measures used. |

Supplementary Table 2. Results of the literature search replications.

| | | Number of records retrieved | | | Percentage difference | | | Speculated cause of discrepancy | |
|---|---|---|---|---|---|---|---|---|---|
| **Review author** | **Database** | **Original reviewers** | **Replicator 1** | **Replicator 2** | **Reviewers & Replicator 1** | **Reviewers & Replicator 2** | **Replicator 1 & Replicator 2** | **Replicator 1** | **Replicator 2** |
| Dohos | PubMed | 8889 | 2921 | 2927 | -67% | -67% | 0% | Reviewer error | Reviewer error |
| | Embase | 11907 | 5722 | 12722 | -52% | 7% | 122% | Different field tags | No discrepancies |
| | CENTRAL | 1778 | 949 | 942 | -47% | -47% | -1% | Reviewer error | Reviewer error |
| | Web of Science | 9538 | 3207 | 2803 | -66% | -71% | -13% | Reviewer error | Reviewer error |
| | Scopus | 14598 | 3673 | 3673 | -75% | -75% | 0% | Reviewer error | Reviewer error |
| Ioannou | PubMed | 912 | 908 | 910 | 0% | 0% | 0% | No discrepancies | No discrepancies |
| | Embase | 1282 | 1281 | 1320 | 0% | 3% | 3% | No discrepancies | No discrepancies |
| | CINAHL | 216 | 169 | 178 | -22% | -18% | 5% | Default settings | Default settings |
| | PsycINFO | 1111 | 1097 | 973 | -1% | -12% | -11% | No discrepancies | Different platform |
| | CENTRAL | 248 | 278 | 241 | 12% | -3% | -13% | Different start date | No discrepancies |
| Yekeduz | MEDLINE | 3412 | 3940 | 3977 | 15% | 17% | 1% | Unreported limit | Unreported limit |
| Zhou | CINAHL | 29 | 24 | 22 | -17% | -24% | -8% | Default settings | Default settings |
| | CENTRAL | 155 | 141 | 157 | -9% | 1% | 11% | No discrepancies | No discrepancies |
| | Embase | 56 | 69 | 70 | 23% | 25% | 1% | Unknown | Unknown |
| | ProQuest | 10 | 71 | 32 | 610% | 220% | -55% | Different resources searched | Different resources searched |
| | PubMed | 55 | 36 | 36 | -35% | -35% | 0% | Platform update | Platform update |
| | Scopus | 137 | 101 | 101 | -26% | -26% | 0% | Unknown | Unknown |
| | Web of Science | 41 | 2209 | 100 | 5288% | 144% | -95% | Incomplete syntax | Incomplete syntax |
| Minozzi | CDAG Register | 5 | NA | NA | NA | NA | NA | NA | NA |
| | CENTRAL | 87 | 245 | 243 | 182% | 179% | -1% | Unknown | Unknown |
| | MEDLINE | 74 | 75 | 75 | 1% | 1% | 0% | No discrepancies | No discrepancies |
| | Embase | 97 | 96 | 90 | -1% | -7% | -6% | No discrepancies | No discrepancies |
| | CINAHL | 248 | 317 | 257 | 28% | 4% | -19% | Different sources | No discrepancies |

| | | | | | | | | | |
|---|---|---|---|---|---|---|---|---|---|
| | PsycINFO | 34 | 37 | 37 | 9% | 9% | 0% | No discrepancies | No discrepancies |
| | Web of Science | 88 | 88 | 91 | 0% | 3% | 3% | No discrepancies | No discrepancies |
| Wang | PubMed | 1980 | 2114 | 2116 | 7% | 7% | 0% | No discrepancies | No discrepancies |
| | Embase | 1408 | 9759 | 1311 | 593% | -7% | -87% | Did not fix errors | No discrepancies |
| | CENTRAL | 42 | 87 | 74 | 107% | 76% | -15% | Unknown | Unknown |
| AlAnouti | MEDLINE | 854 | 841 | 841 | -2% | -2% | 0% | No discrepancies | No discrepancies |
| | PubMed | 1028 | 991 | 991 | -4% | -4% | 0% | No discrepancies | No discrepancies |
| | CINAHL | 852 | 755 | 785 | -11% | -8% | 4% | Unknown | No discrepancies |
| | Embase | 941 | 909 | 918 | -3% | -2% | 1% | No discrepancies | No discrepancies |
| | CENTRAL | 900 | 857 | 951 | -5% | 6% | 11% | No discrepancies | No discrepancies |
| Goldberg | PubMed, CINAHL & PsycINFO | 5389 | 6910 | 4852 | 28% | -10% | -30% | Unknown | Unknown |
| | PubMed | NA | 3130 | 2965 | NA | NA | -5% | NA | NA |
| | CINAHL | NA | 743 | 506 | NA | NA | -32% | NA | NA |
| | PsycINFO | NA | 3037 | 1381 | NA | NA | -55% | NA | NA |
| | Web of Science | 2167 | 2334 | 2620 | 8% | 21% | 12% | No discrepancies | Different resources searched |
| | Scopus | 6650 | 5056 | 5046 | -24% | -24% | 0% | Different field tags | Different field tags |
| | CENTRAL | 380 | 390 | 373 | 3% | -2% | -4% | No discrepancies | No discrepancies |
| Jakubczyk | PubMed | 473 | 634 | 635 | 34% | 34% | 0% | Undisclosed limits | Undisclosed limits |
| | Embase | 286 | 341 | 335 | 19% | 17% | -2% | Undisclosed limits | Undisclosed limits |
| | PubMed | 98 | 98 | 98 | 0% | 0% | 0% | No discrepancies | No discrepancies |
| Gingold-Belfer | MEDLINE | 96 | 98 | 98 | 2% | 2% | 0% | No discrepancies | No discrepancies |
| | Embase | 453 | 434 | 455 | -4% | 0% | 5% | No discrepancies | No discrepancies |
| | Web of Science | 61 | 102 | 98 | 67% | 61% | -4% | Different resources searched | Different resources searched |
| | CENTRAL | 50 | 26 | 24 | -48% | -52% | -8% | Unknown | Unknown |

Supplementary Table 3. Investigations of the 13 different screening decisions and the suspected reasons for their cause.

| Review | Study | Original reviewers | Blinded replicators | Discrepancy reason | Discrepancy details | Unblinded replicators | Changed decision |
|---|---|---|---|---|---|---|---|
| AlAnouti 2020 | Yin 2016 | Exclude | Include | Coordinator error | The REPRISE study coordinator missed guidance in the review's results section which outlined that they excluded this study due to the follow up period being too long (i.e., the eligible time frames for the outcome was not specified in the methods section). | Exclude | Yes |
| Dohos 2020 | Candy 1995 | Include | Exclude | Replicator error | Both replicators missed that the study authors did report the outcome of interest. That is, how many participants responded to the 12-weeks prednisolone treatment, and of them, the number of people who stayed in remission after receiving azathioprine or placebo. | Include | Yes |
| Dohos 2020 | O'Donoghue 1978 | Include | Exclude | Lack of content expertise | This study included a group which received both an immunomodulator and anti-inflammatory drugs. The original reviewers did not provide specific guidance as to whether anti-inflammatory drugs are an acceptable co-intervention for IM monotherapy. Given the lack of guidance and content expertise, the blinded replicators decided to exclude this study. After reading the review report and other relevant literature, we decided that this was likely considered an acceptable co-intervention. | Include | Yes |
| Ioannou 2020 | Elsenga 1982 | Include | Exclude | Incomplete eligibility criteria | The original reviewers explicitly stated that studies would only be included if they followed DSM criteria. It was not specified in the primary study whether study participants had depression as defined according to DSM criteria. Consequently, the replicators decided to exclude this study due to a lack of information (note that an attempt was made to contact the study authors which was unsuccessful). When the original reviewers were asked about the eligibility assessment of this study, they stated via email that they decided to include the study based on information contained in the study author's dissertation. A clinical psychologist was also consulted to evaluate the similarity of the diagnostic criteria used by the study authors in comparison to the DSM. Their view was also that the two criteria (Elsenga's and DSM III) were sufficiently similar. | Include | Yes |
| Ioannou 2020 | Kragh 2017a | Include | Exclude | Unclear eligibility criteria | The original reviewers explicitly stated that studies would only be included if they used DSM criteria to diagnose depression. The study authors stated that depression was diagnosed according to ICD-10 | Exclude | No |

| Review | Study | Original reviewers | Blinded replicators | Discrepancy reason | Discrepancy details | Unblinded replicators | Changed decision |
|---|---|---|---|---|---|---|---|
| | | | | | criteria. Therefore, the blinded replicators excluded this study. After unblinding, the replicators noted that this study was included in the original review. Following this discovery the replicators looked at the two diagnostic criteria, as well as consulted with a clinical psychologist, and maintained their view that ICD-10 and DSM III criteria were too dissimilar (as well as at direct odds with the instructions given) and so did not include this study in the unblinded replication either. Consequently, it appears that either the original reviewers did not follow their own eligibility criteria, or the allowance of the ICD-10 criteria was an undeclared change to the eligibility criteria. | | |
| Ioannou 2020 | Wu 2009 | Include | Exclude | Incomplete eligibility criteria | The intervention in this study consisted of sleep deprivation and sleep phase advance. While the original reviewers stated that light therapy was considered an acceptable co-intervention, they did not provide guidance on the acceptability of other chronotherapies (e.g., sleep phase advance, sleep time stabilisation). Given the lack of information, the blinded replicators decided to exclude this study. After being unblinded and reading the limitations section in the discussion section of the review report it appears that the original reviewers also considered sleep phase advance (as well as sleep time stabilisation) to be an acceptable chronotherapeutic co-intervention in addition to 'light therapy'. | Include | Yes |
| Wang 2020 | Akiyama 1994 | Exclude | Include | Coordinator error | The REPRISE study coordinator failed to notice that the original reviewers indicated that they would exclude studies with a Newcastle-Ottawa Score less than six from the meta-analysis. | Exclude | Yes |
| Wang 2020 | Fujita 2003 | Exclude | Include | Coordinator error | The REPRISE study coordinator failed to notice that the original reviewers indicated that they would exclude studies with a Newcastle-Ottawa Score less than six from the meta-analysis. | Exclude | Yes |
| Wang 2020 | Koterazawa 2019 | Exclude | Include | Coordinator error | The REPRISE study coordinator failed to notice that the original reviewers indicated that they would exclude studies with a Newcastle-Ottawa Score less than six from the meta-analysis. | Exclude | Yes |
| Yekeduz 2020 | Lee 2020b | Exclude | Include | Replicator error | The replicators failed to notice that this study's population overlapped with the population from the Lee 2020a report (which was also included in the original review). The replicators assume Lee 2020a was included in | Exclude | Yes |

| Review | Study | Original reviewers | Blinded replicators | Discrepancy reason | Discrepancy details | Unblinded replicators | Changed decision |
|---|---|---|---|---|---|---|---|
| | | | | | place of this study as it reports on a bigger sample size and a broader cohort (i.e., not just haematological malignancies). | | |
| Zhou 2021 | Hoseini 2016 | Exclude | Include | Unclear | The original reviewers state in the narrative synthesis section (3.8) that they were unable to retrieve sufficient information for inclusion of this study. However, to the replicators, all required information appears to have been reported in the study's abstract. Therefore, it is unclear to us exactly why this study was not included by the original reviewers. | Include | No |
| Zhou 2021 | Minatel 2009 | Include | Exclude | Ambiguous eligibility criteria | The REPRISE study coordinator advised the blinded replicators that participants with "lower limb diabetic foot ulcers" were eligible for the index meta-analysis as this was stated in the eligibility criteria section in the methods section. The replicators also noted that no explicit guidance was provided regarding whether the original reviewers were interested in diabetic (neuropathic) ulcers and/or other ulcers (e.g., ulcers due to venous or arterial insufficiency) in diabetic populations. Given the presence of the term 'diabetic foot ulcer' in the criterion, the blinded replicators decided to interpret "lower limb diabetic foot ulcers" as neuropathic ulcers occurring on the foot (i.e., ulcers distal to the ankle), not leg, and hence excluded this study as it included participants with both mixed ulcers, as well as ulcers above the ankle. Upon reading the original review's supplementary material after unblinding, we noted that the original reviewers stated that "lower *limb* wounds/ulcers" were deemed eligible. After seeing this phrasing, we reasoned that the original reviewers likely intended that the phrase "lower limb diabetic foot ulcers" refer broadly to diabetic ulcers, *as well as* mixed (e.g., venous and arterial) ulcers in people with Type II diabetes, occurring anywhere on the lower extremity. However, we were not able to confirm this updated interpretation of the eligibility criteria with the original reviewers. | Include | Yes |
| Zhou 2021 | Zhang 2013 | Include | Exclude | Unclear eligibility criteria | The original reviewers explicitly stated in the detailed eligibility criteria specified in the supplement that articles in languages other than English were ineligible. As this study is written in Chinese, it was excluded by the replicators in both phases. Consequently, it appears that either the original reviewers did not follow their own eligibility criteria, or that the inclusion of foreign-language articles was an undeclared change to the eligibility criteria. We did not change our decision as both the | Exclude | No |

| Review | Study | Original reviewers | Blinded replicators | Discrepancy reason | Discrepancy details | Unblinded replicators | Changed decision |
|---|---|---|---|---|---|---|---|
| | | | | | PROSPERO entry and report say that they did not include non-English language studies. | | |

Supplementary Table 4. Investigations of the reasons for the data extraction discrepancies.

| Review | Study | EE type | Original reviewers | | Blinded replicators | | Unblinded replicators | | Discrepancy reason | Discrepancy details |
|---|---|---|---|---|---|---|---|---|---|---|
| | | | EE | 95% CI | EE | 95% CI | EE | 95% CI | | |
| AlAnouti 2020 | Wongwiwat thananukit 2013 | MD | 2.04 | (-31.00, 35.08) | 2.04 | (-29.88, 33.96) | NA | NA | Reviewer error | The original reviewers appear to have not noticed that the study authors imputed data for the 2 people in each group who withdrew prior to the Week 8 mark. |
| Dohos 2020 | Hawthorne 1992 | Risk ratio | 1.39 | (0.85, 2.26) | 1.62 | (0.95, 2.75) | 1.52 | (0.90, 2.57) | Incomplete data selection methods | No guidance was provided in the review report on which timepoint to choose. The blinded replicators chose the result reported in text which represents the relapse rate at eight months (i.e., the time prior to the first person dropping out). Whereas, after being unblinded, and in consultation with a biostatistician, the replicators and decided to use the data at 12 months which required removing the two censored patients. Differences between both replicators' results and the original reviewers' results likely relate to differences in the assumption about what has happened to the two people who dropped out. We assumed they didn't relapse. The review authors may not have assumed this. |
| Dohos 2020 | O'Donoghue 1978 | Risk ratio | 2.44 | (0.90, 6.67) | NA | NA | 7.56 | (1.04, 54.91) | Incomplete data selection methods | No guidance was provided on how to account for patients who dropped out. Differences between the replicators and original reviewers likely relate to the replicators assuming the five participants who exited the trial for 'reasons other than relapse' didn't relapse. The review authors may not have assumed this. |
| Dohos 2020 | Vilien 2004 | Risk ratio | 2.49 | (0.82, 7.55) | 3.47 | (0.89, 13.51) | NA | NA | Incomplete data selection methods | No guidance was provided on how to account for patients who dropped out. Differences between the replicators and original reviewers likely relate to the replicators assuming that the person who exited the trial didn't relapse. The review authors may not have assumed this. |
| Gingold-Belfer 2020 | Perri 2001 | Odds ratio | 1.64 | (0.85, 3.17) | 1.64 | (0.75, 3.62) | NA | NA | Reviewer error | It appears that the original reviewers have double counted the control group (60/90) when calculating the odds ratio. |

| Review | Study | EE type | Original reviewers | | Blinded replicators | | Unblinded replicators | | Discrepancy reason | Discrepancy details |
|---|---|---|---|---|---|---|---|---|---|---|
| | | | EE | 95% CI | EE | 95% CI | EE | 95% CI | | |
| Goldberg 2020 | Gasser 2014 | SMD | 2.22 | (0.91, 3.53) | 0.09 | (-1.24, 1.42) | NA | NA | Incomplete data selection methods | No specific guidance was provided on which outcome measure to choose when multiple measures were available. Therefore, it is very likely that differences occurred due to the choice of outcome used to serve as the surrogate for 'targeted symptoms'. The replicators decided to choose the outcome scale they felt was most applicable to the population's condition of interest (e.g., depression-only scale for populations with depression, combined depression and anxiety scale for populations with both anxiety and depression) as well as to use results from the same scale from each study when multiple scales were reported. The original reviewers stated upon request for clarification that they no longer have the data for the review, so we are unable to confirm that this was the cause of the discrepancy. |
| Goldberg 2020 | Griffiths 2016 | SMD | 0.86 | (0.24, 1.48) | 0.55 | (-0.01, 1.11) | NA | NA | Incomplete data selection methods | As per Gasser 2014 comment. |
| Goldberg 2020 | Grob 2011 | SMD | 0.94 | (-0.06, 1.94) | 1.21 | (-0.04, 2.46) | NA | NA | Incomplete data selection methods | As per Gasser 2014 comment. |
| Goldberg 2020 | Palhano-Fontes 2019 | SMD | 1.20 | (0.52, 1.88) | 1.77 | (0.90, 2.64) | NA | NA | Incomplete data selection methods | As per Gasser 2014 comment. |
| Goldberg 2020 | Ross 2016 | SMD | 0.98 | (0.27, 1.69) | 0.61 | (-0.14, 1.35) | NA | NA | Incomplete data selection methods | As per Gasser 2014 comment. |
| Ioannou 2020 | Benedetti 1997 | SMD | -0.51 | (-1.78, 0.77) | -0.61 | (-1.90, 0.67) | NA | NA | Reviewer error | It appears that the original reviewers incorrectly extracted the Day 14 results rather than the Day 7 results. The review report specifies that the outcome of interest is the depression scores within one week of starting treatment. Study authors state that participants commenced sleep deprivation on Day 6. |

| Review | Study | EE type | Original reviewers EE | Original reviewers 95% CI | Blinded replicators EE | Blinded replicators 95% CI | Unblinded replicators EE | Unblinded replicators 95% CI | Discrepancy reason | Discrepancy details |
|---|---|---|---|---|---|---|---|---|---|---|
| Ioannou 2020 | Kunderman n 2008 | SMD | -0.17 | (-1.07, 0.73) | -0.17 | (-1.08, 0.73) | -0.17 | (-1.08, 0.73) | Data extraction from plot | The difference is due to a small variation in the value of the standard deviation of the control group that was estimated from Figure 2. |
| Wang 2020 | Fujita 1995 | HR | 1.13 | (0.67, 1.92) | 1.19 | (0.77, 1.83) | 1.19 | (0.77, 1.83) | Data extraction from plot | The difference is due to a combination of the manual extraction of data from reported Kaplan-Meier (KM) curves and the estimation of the hazard ratio and standard error from the data via Guyot's algorithm. We noted during calibration testing that extracting more datapoints from KM curves, and the addition of other pertinent data into the algorithm (e.g., data from risk tables), resulted in more accurate estimates. |
| Wang 2020 | Igaki 2004 | | 1.29 | (0.77, 2.15) | 1.25 | (0.80, 1.96) | 1.25 | (0.80, 1.96) | Data extraction from plot | As per Fujita 1995 comment. |
| Wang 2020 | Li 2012 | | 0.88 | (0.63, 1.23) | 0.88 | (0.62, 1.26) | 0.88 | (0.62, 1.26) | Data extraction from plot | As per Fujita 1995 comment. |
| Wang 2020 | Shim 2010 | | 1.35 | (0.71, 2.58) | 1.15 | (0.64, 2.07) | 1.15 | (0.64, 2.07) | Data extraction from plot | As per Fujita 1995 comment. |
| Wang 2020 | Tabira 1999 | | 1.20 | (0.65, 2.20) | 1.52 | (0.89, 2.57) | 1.52 | (0.89, 2.57) | Data extraction from plot | As per Fujita 1995 comment. |
| Wang 2020 | Zhang 2008 | | 1.31 | (0.82, 2.10) | 1.30 | (0.86, 1.96) | 1.30 | (0.86, 1.96) | Data extraction from plot | As per Fujita 1995 comment. |
| Zhou 2021 | Naidu 2005 | SMD | 4.17 | (2.45, 5.89) | 2.92 | (1.40, 4.43) | NA | NA | Unclear | No specific guidance was provided on which timepoint to choose when outcome data from multiple timepoints were available. Consequently, the blinded replicators chose the Week 2 result to ensure consistency with other included studies (which all provided outcome data at Week 2). After this discrepancy was discovered following unblinding, Hedge's g was calculated for all available |

| Review | Study | EE type | Original reviewers | | Blinded replicators | | Unblinded replicators | | Discrepancy reason | Discrepancy details |
|---|---|---|---|---|---|---|---|---|---|---|
| | | | EE | 95% CI | EE | 95% CI | EE | 95% CI | | |
| | | | | | | | | | | timepoints to determine if a different timepoint was chosen by the original reviewers. However, we were unable to reconstruct the reported value from the review. The replicators tried to confirm with the original reviewers but received no response. |

Supplementary Table 5. Percentage differences between the original review's and replicators' summary estimate and confidence interval widths. Note: Ratio measures were log-transformed (natural log) prior to calculation of the percentage difference.

| | | | | Percentage difference | |
|---|---|---|---|---|---|
| **Review** | **Analysis** | **Summary estimate** | **95% CI** | **Summary estimate** | **CI width** |
| Dohos 2020 | Original (report) | 1.85 | (1.44, 2.38) | NA | NA |
| | Replicator 1 (blinded) | 1.844 | (1.336, 2.545) | -1% | 28% |
| | Replicator 2 (blinded) | 1.844 | (1.337, 2.543) | -1% | 28% |
| | Replicator 1 (unblinded) | 1.943 | (1.487, 2.537) | 8% | 6% |
| | Replicator 2 (unblinded) | 1.943 | (1.488, 2.537) | 8% | 6% |
| Ioannou 2020 | Original (report) | -0.29 | (-0.84, 0.25) | NA | NA |
| | Replicator 1 (blinded) | -0.319 | (-1.058, 0.419) | 10% | 36% |
| | Replicator 2 (blinded) | -0.332 | (-1.022, 0.358) | 14% | 27% |
| | Replicator 1 (unblinded) | -0.267 | (-0.962, 0.428) | -8% | 28% |
| | Replicator 2 (unblinded) | -0.276 | (-0.961, 0.409) | -5% | 26% |
| Yekeduz 2020 | Original (report) | 1.85 | (1.26, 2.71) | NA | NA |
| | Replicator 1 (blinded) | 1.533 | (1.119, 2.10) | -31% | -18% |
| | Replicator 2 (blinded) | 1.533 | (1.118, 2.10) | -31% | -18% |
| | Replicator 1 (unblinded) | 1.846 | (1.26, 2.705) | 0% | 0% |
| | Replicator 2 (unblinded) | 1.847 | (1.26, 2.705) | 0% | 0% |
| Zhou 2021 | Original (report) | 2.81 | (1.14, 4.48) | NA | NA |
| | Replicator 1 (blinded) | 3.621 | (2.311, 4.93) | 29% | -22% |
| | Replicator 2 (blinded) | 3.62 | (2.326, 4.913) | 29% | -23% |
| | Replicator 1 (unblinded) | 3.381 | (2.211, 4.551) | 20% | -30% |
| | Replicator 2 (unblinded) | 3.376 | (2.224, 4.528) | 20% | -31% |
| Minozzi 2020 | Original (report) | 0.66 | (0.37, 1.2) | NA | NA |
| | Replicator 1 (blinded) | 0.662 | (0.366, 1.197) | -1% | 1% |
| | Replicator 2 (blinded) | 0.661 | (0.367, 1.193) | 0% | 0% |
| | Replicator 1 (unblinded) | 0.662 | (0.366, 1.197) | -1% | 1% |
| | Replicator 2 (unblinded) | 0.661 | (0.367, 1.193) | 0% | 0% |
| Wang 2020 | Original (report) | 1.05 | (0.9, 1.21) | NA | NA |
| | Replicator 1 (blinded) | 1.055 | (0.944, 1.182) | 10% | -24% |
| | Replicator 2 (blinded) | 1.058 | (0.946, 1.183) | 16% | -24% |
| | Replicator 1 (unblinded) | 1.069 | (0.929, 1.23) | 37% | -5% |
| | Replicator 2 (unblinded) | 1.068 | (0.929, 1.229) | 35% | -5% |
| AlAnouti 2020 | Original (report) | 30.67 | (4.89, 56.45) | NA | NA |
| | Replicator 1 (blinded) | 0.368 | (-9.185, 9.922) | -99% | -63% |
| | Replicator 2 (blinded) | 0.46 | (-9.048, 9.969) | -99% | -63% |
| | Replicator 1 (unblinded) | 29.473 | (4.236, 54.71) | -4% | -2% |
| | Replicator 2 (unblinded) | 30.022 | (4.937, 55.107) | -2% | -3% |

Supplementary Table 5 (cont). Percentage differences between the original review's and replicators' summary estimate and confidence interval widths. Note: Ratio measures were log-transformed (natural log) prior to calculation of the percentage difference.

| | | | | Percentage difference | |
|---|---|---|---|---|---|
| **Review** | **Analysis** | **Summary estimate** | **95% CI** | **Summary estimate** | **CI width** |
| Goldberg 2020 | Original (report) | 1.08 | (0.74, 1.43) | NA | NA |
| | Replicator 1 (blinded) | 0.839 | (0.308, 1.37) | -22% | 54% |
| | Replicator 2 (blinded) | 0.842 | (0.31, 1.375) | -22% | 54% |
| | Replicator 1 (unblinded) | 0.839 | (0.308, 1.37) | -22% | 54% |
| | Replicator 2 (unblinded) | 0.842 | (0.31, 1.375) | -22% | 54% |
| Jakubczyk 2020 | Original (report) | 2.7 | (0.06, 5.34) | NA | NA |
| | Replicator 1 (blinded) | 2.683 | (0.082, 5.284) | -1% | -1% |
| | Replicator 2 (blinded) | 2.696 | (0.058, 5.335) | 0% | 0% |
| | Replicator 1 (unblinded) | 2.683 | (0.082, 5.284) | -1% | -1% |
| | Replicator 2 (unblinded) | 2.696 | (0.058, 5.335) | 0% | 0% |
| Gingold-Belfer 2020 | Original (report) | 0.89 | (0.44, 1.79) | NA | NA |
| | Replicator 1 (blinded) | 0.869 | (0.43, 1.759) | 20% | 0% |
| | Replicator 2 (blinded) | 0.87 | (0.43, 1.759) | 20% | 0% |
| | Replicator 1 (unblinded) | 0.869 | (0.43, 1.759) | 20% | 0% |
| | Replicator 2 (unblinded) | 0.87 | (0.43, 1.759) | 20% | 0% |

Supplementary Table 6. Discordance between P values for the test for overall effect from the original review's and replicators' meta-analyses.

| Review | Analysis | P value (overall) | P < 0.01 | 0.01 ≤ P < 0.05 | 0.05 ≤ P < 0.1 | P ≥ 0.1 |
|---|---|---|---|---|---|---|
| Dohos 2020 | Original (report) | 0 | Yes | - | - | - |
| | Replicator 1 (blinded) | 0 | Yes | - | - | - |
| | Replicator 2 (blinded) | 0 | Yes | - | - | - |
| | Replicator 1 (unblinded) | 0 | Yes | - | - | - |
| | Replicator 2 (unblinded) | 0 | Yes | - | - | - |
| Ioannou 2020 | Original (report) | 0.29 | - | - | - | Yes |
| | Replicator 1 (blinded) | 0.397 | - | - | - | Yes |
| | Replicator 2 (blinded) | 0.346 | - | - | - | Yes |
| | Replicator 1 (unblinded) | 0.452 | - | - | - | Yes |
| | Replicator 2 (unblinded) | 0.43 | - | - | - | Yes |
| Yekeduz 2020 | Original (report) | 0 | Yes | - | - | - |
| | Replicator 1 (blinded) | 0.008 | Yes | - | - | - |
| | Replicator 2 (blinded) | 0.008 | Yes | - | - | - |
| | Replicator 1 (unblinded) | 0.002 | Yes | - | - | - |
| | Replicator 2 (unblinded) | 0.002 | Yes | - | - | - |
| Zhou 2021 | Original (report) | 0 | Yes | - | - | - |
| | Replicator 1 (blinded) | 0 | Yes | - | - | - |
| | Replicator 2 (blinded) | 0 | Yes | - | - | - |
| | Replicator 1 (unblinded) | 0 | Yes | - | - | - |
| | Replicator 2 (unblinded) | 0 | Yes | - | - | - |
| Minozzi 2020 | Original (report) | 0.17 | - | - | - | Yes |
| | Replicator 1 (blinded) | 0.172 | - | - | - | Yes |
| | Replicator 2 (blinded) | 0.169 | - | - | - | Yes |
| | Replicator 1 (unblinded) | 0.172 | - | - | - | Yes |
| | Replicator 2 (unblinded) | 0.169 | - | - | - | Yes |
| Wang 2020 | Original (report) | 0.56 | - | - | - | Yes |
| | Replicator 1 (blinded) | 0.342 | - | - | - | Yes |
| | Replicator 2 (blinded) | 0.326 | - | - | - | Yes |
| | Replicator 1 (unblinded) | 0.352 | - | - | - | Yes |
| | Replicator 2 (unblinded) | 0.356 | - | - | - | Yes |
| AlAnouti 2020 | Original (report) | 0.02 | - | Yes | - | - |
| | Replicator 1 (blinded) | 0.94 | - | - | - | Yes |
| | Replicator 2 (blinded) | 0.925 | - | - | - | Yes |
| | Replicator 1 (unblinded) | 0.022 | - | Yes | - | - |
| | Replicator 2 (unblinded) | 0.019 | - | Yes | - | - |

Supplementary Table 6 (cont). Discordance between P values for the test for overall effect from the original review's and replicators' meta-analyses.

| Review | Analysis | P value (overall) | P < 0.01 | 0.01 ≤ P < 0.05 | 0.05 ≤ P < 0.1 | P ≥ 0.1 |
|---|---|---|---|---|---|---|
| Goldberg 2020 | Original (estimated in R) | 0 | Yes | - | - | - |
| | Replicator 1 (blinded) | 0.002 | Yes | - | - | - |
| | Replicator 2 (blinded) | 0.002 | Yes | - | - | - |
| | Replicator 1 (unblinded) | 0.002 | Yes | - | - | - |
| | Replicator 2 (unblinded) | 0.002 | Yes | - | - | - |
| Jakubczyk 2020 | Original (report) | 0.045 | - | Yes | - | - |
| | Replicator 1 (blinded) | 0.043 | - | Yes | - | - |
| | Replicator 2 (blinded) | 0.045 | - | Yes | - | - |
| | Replicator 1 (unblinded) | 0.043 | - | Yes | - | - |
| | Replicator 2 (unblinded) | 0.045 | - | Yes | - | - |
| Gingold-Belfer 2020 | Original (report) | 0.73 | - | - | - | Yes |
| | Replicator 1 (blinded) | 0.698 | - | - | - | Yes |
| | Replicator 2 (blinded) | 0.698 | - | - | - | Yes |
| | Replicator 1 (unblinded) | 0.698 | - | - | - | Yes |
| | Replicator 2 (unblinded) | 0.698 | - | - | - | Yes |

Supplementary Table 7. Factors associated with the eight blinded replicated meta-analyses classified as not fully replicable. (Note: '+' refers to suspected minor contributors to replication failure and '++' refers to suspected major contributors to replication failure.)

| | Replicator-related | | | Error | Reporting | | | Other | | | |
|---|---|---|---|---|---|---|---|---|---|---|---|
| | Coordinator error | Replicator error | Lack of content expertise | Reviewer error | Incomplete eligibility criteria | Ambiguous eligibility criteria | Incomplete data selection methods | Extraction of data from plots | Software-related differences | Unavailable data | Unknown reasons |
| AlAnouti 2020 | ++ | - | - | + | - | - | - | - | - | - | - |
| Dohos 2020 | - | ++ | ++ | - | - | - | + | - | - | - | - |
| Gingold-Belfer 2020 | - | - | - | ++ | - | - | - | - | - | - | - |
| Goldberg 2020 | - | - | - | - | - | - | ++ | - | - | - | - |
| Ioannou 2020 | - | - | - | ++ | ++ | ++ | - | + | - | ++ | - |
| Wang 2020 | ++ | - | - | - | - | - | - | ++ | + | - | - |
| Yekeduz 2020 | - | ++ | - | - | - | - | - | - | + | - | - |
| Zhou 2021 | - | - | - | - | - | ++ | - | - | + | - | ++ |

Supplementary Table 8. Factors associated with the five unblinded replicated meta-analyses classified as not fully replicable. (Note: '+' refers to suspected minor contributors to replication failure and '++' refers to suspected major contributors to replication failure.)

| | Replicator-related | | | Error | Reporting | | | Other | | | |
|---|---|---|---|---|---|---|---|---|---|---|---|
| | Coordinator error | Replicator error | Lack of content expertise | Reviewer data extraction error | Incomplete eligibility criteria | Ambiguous eligibility criteria | Incomplete data selection methods | Extraction of data from plots | Software-related differences | Unavailable data | Unknown reasons |
| Gingold-Belfer 2020 | - | - | - | ++ | - | - | - | - | - | - | - |
| Goldberg 2020 | - | - | - | - | - | - | ++ | - | - | - | - |
| Ioannou 2020 | - | - | - | ++ | - | ++ | - | + | - | - | - |
| Wang 2020 | - | - | - | - | - | - | - | ++ | + | - | - |
| Zhou 2021 | - | - | - | - | - | ++ | - | - | + | - | ++ |

### SUPPLEMENTARY FIGURES

Supplementary Figure 1. Scatter plot of the number of records retrieved by the original reviewers (square symbols) and the first (plus signs) and second (cross signs) replicators by database. Note: The y-axis is presented on a logarithmic scale (base 10).

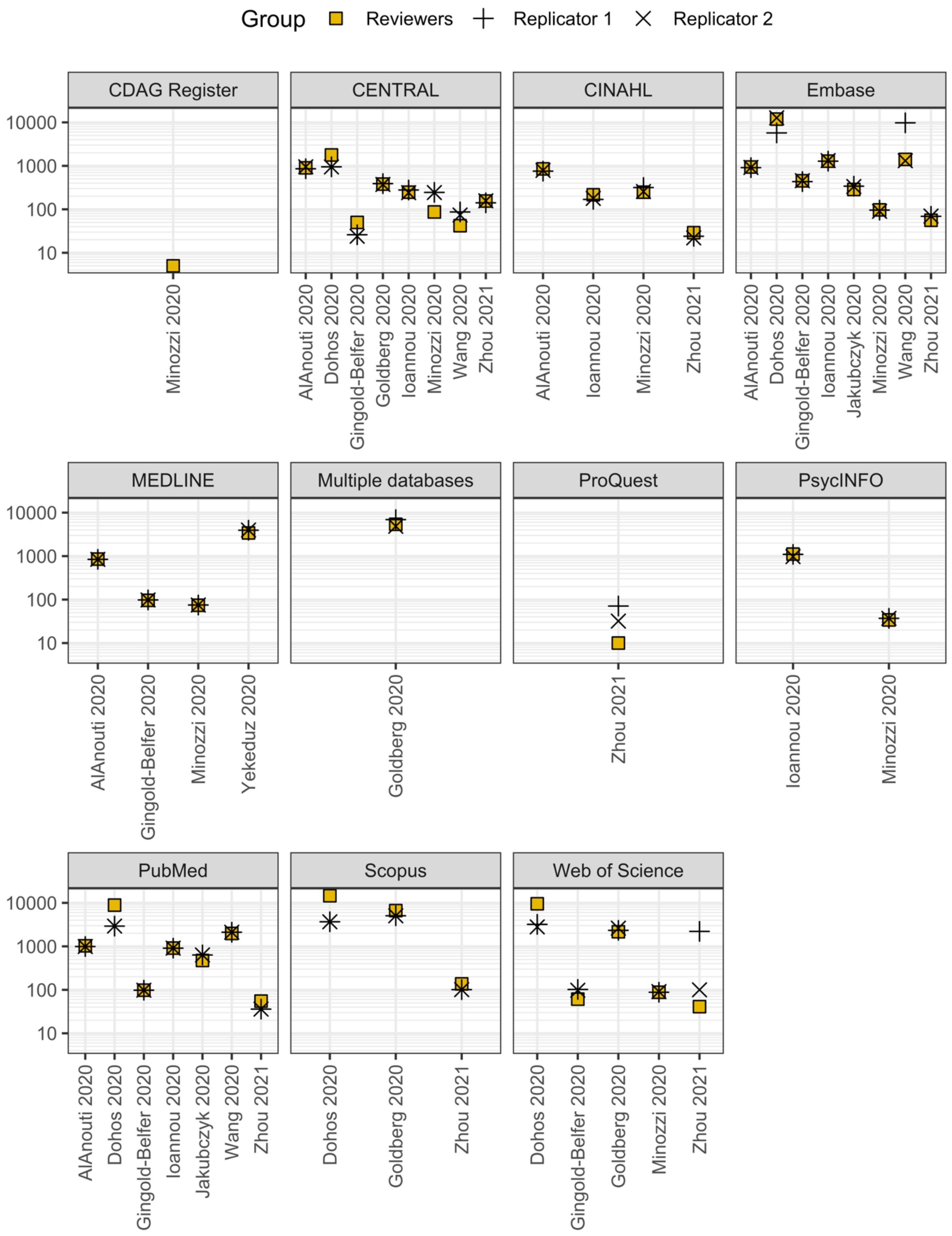

Supplementary Figure 2. Results of the original and replicated index meta-analyses for the review by AlAnouti et al. (2020) [5]. (Green shade: Met the criteria for ‘fully replicable’. Red shade: Met the criteria for not ‘fully replicable’. Black shade: Met the criteria for not ‘fully replicable’ and ‘meaningfully’ different.)

| | Intervention | | | Control | | | | | Screening discrepancy | Data discrepancy | Reason for discrepancy |
|---|---|---|---|---|---|---|---|---|---|---|---|
| | Mean | SD | Total | Mean | SD | Total | MD | 95% CI | | | |
| Original reviewers | | | | | | | | | | | |
| Farag et al. (2019) | 233.8 | 97.0 | 24 | 158.6 | 35.4 | 25 | 75.20 | (33.99, 116.41) | - | - | - |
| Wongwiwatthananukit (2013) | 137.8 | 53.5 | 28 | 135.8 | 71.4 | 28 | 2.04 | (-31.00, 35.08) | - | - | - |
| Yin et al. (2016) | NI | NI | NI | NI | NI | NI | NI | NI | - | - | - |
| [Fixed effect model] | - | - | - | - | - | - | 30.67 | (4.89, 56.45) | | | |
| Replicators (Blinded) | | | | | | | | | | | |
| Farag et al. (2019) | 233.8 | 97.0 | 24 | 158.6 | 35.4 | 25 | 75.20 | (33.99, 116.41) | No | No | - |
| Wongwiwatthananukit (2013) | 137.8 | 53.5 | 30 | 135.8 | 71.4 | 30 | 2.04 | (-29.88, 33.96) | No | Yes | Reviewer extraction error |
| Yin et al. (2016) | 250.4 | 36.3 | 61 | 254.9 | 19.5 | 62 | -4.43 | (-14.74, 5.88) | Yes | - | Coordinator extraction error |
| [Fixed effect model (R1)] | - | - | - | - | - | - | 0.37 | (-9.19, 9.92) | | | |
| [Fixed effect model (R2)] | - | - | - | - | - | - | 0.46 | (-9.05, 9.97) | | | |
| Replicators (Unblinded) | | | | | | | | | | | |
| Farag et al. (2019) | 233.8 | 97.0 | 24 | 158.6 | 35.4 | 25 | 75.20 | (33.99, 116.41) | No | No | - |
| Wongwiwatthananukit (2013) | 137.8 | 53.5 | 30 | 135.8 | 71.4 | 30 | 2.04 | (-29.88, 33.96) | No | Yes | Reviewer extraction error |
| Yin et al. (2016) | NI | NI | NI | NI | NI | NI | NI | NI | No | - | - |
| [Fixed effect model (R1)] | - | - | - | - | - | - | 29.47 | (4.24, 54.71) | | | |
| [Fixed effect model (R2)] | - | - | - | - | - | - | 30.02 | (4.94, 55.11) | | | |

Supplementary Figure 3. Results of the original and replicated index meta-analyses for the review by Dohos et al (2021) [6]. (Green shade: Met the criteria for 'fully replicable'. Red shade: Met the criteria for not 'fully replicable'. Black shade: Met the criteria for not 'fully replicable' and 'meaningfully' different.)

| | Intervention | | Control | | | | Screening discrepancy | Data discrepancy | Reason for discrepancy |
|---|---|---|---|---|---|---|---|---|---|
| | Events | Total | Events | Total | RR | 95% CI | | | |
| Original reviewers | | | | | | | | | |
| Feagan et al. (2000) | 22 | 36 | 14 | 40 | 1.75 | (1.06, 2.87) | - | - | - |
| Wenzl et al. (2014) | 8 | 26 | 4 | 26 | 2.00 | (0.69, 5.83) | - | - | - |
| Candy et al. (1995) | 17 | 19 | 10 | 24 | 2.15 | (1.31, 3.53) | - | - | - |
| O'Donoghue et al. (1978) | 11 | 27 | 4 | 24 | 2.44 | (0.90, 6.67) | - | - | - |
| Vilien et al. (2004) | 8 | 15 | 3 | 14 | 2.49 | (0.82, 7.55) | - | - | - |
| Lémann et al. (2005) | 9 | 43 | 3 | 40 | 2.79 | (0.81, 9.58) | - | - | - |
| Hawthorne et al. (1992) | 20 | 34 | 14 | 33 | 1.39 | (0.85, 2.26) | - | - | - |
| [Random effects model] | - | - | - | - | 1.85 | (1.44, 2.38) | | | |
| Replicators (Blinded) | | | | | | | | | |
| Feagan et al. (2000) | 22 | 36 | 14 | 40 | 1.75 | (1.06, 2.87) | No | No | - |
| Wenzl et al. (2014) | 8 | 26 | 4 | 26 | 2.00 | (0.69, 5.83) | No | No | - |
| Candy et al. (1995) | NI | NI | NI | NI | NI | NI | Yes | - | Replicator screening error |
| O'Donoghue et al. (1978) | NI | NI | NI | NI | NI | NI | Yes | - | Lack of content expertise |
| Vilien et al. (2004) | 8 | 15 | 2 | 13 | 3.47 | (0.89, 13.51) | No | Yes | Incomplete data selection methods |
| Lémann et al. (2005) | 9 | 43 | 3 | 40 | 2.79 | (0.81, 9.58) | No | No | - |
| Hawthorne et al. (1992) | 20 | 34 | 12 | 33 | 1.62 | (0.95, 2.75) | No | Yes | Incomplete data selection methods |
| [Random effects model (R1)] | - | - | - | - | 1.84 | (1.34, 2.55) | | | |
| [Random effects model (R2)] | - | - | - | - | 1.84 | (1.34, 2.54) | | | |
| Replicators (Unblinded) | | | | | | | | | |
| Feagan et al. (2000) | 22 | 36 | 14 | 40 | 1.75 | (1.06, 2.87) | No | No | - |
| Wenzl et al. (2014) | 8 | 26 | 4 | 26 | 2.00 | (0.69, 5.83) | No | No | - |
| Candy et al. (1995) | 17 | 19 | 10 | 24 | 2.15 | (1.31, 3.53) | No | No | - |
| O'Donoghue et al. (1978) | 9 | 25 | 1 | 21 | 7.56 | (1.04, 54.91) | No | Yes | Incomplete data selection methods |
| Vilien et al. (2004) | 8 | 15 | 2 | 13 | 3.47 | (0.89, 13.51) | No | Yes | Incomplete data selection methods |
| Lémann et al. (2005) | 9 | 43 | 3 | 40 | 2.79 | (0.81, 9.58) | No | No | - |
| Hawthorne et al. (1992) | 20 | 34 | 12 | 31 | 1.52 | (0.90, 2.57) | No | Yes | Incomplete data selection methods |
| [Random effects model (R1)] | - | - | - | - | 1.94 | (1.49, 2.54) | | | |
| [Random effects model (R2)] | - | - | - | - | 1.94 | (1.49, 2.54) | | | |

Supplementary Figure 4. Results of the original and replicated index meta-analyses for the review by Gingold-Belfer et al. (2021) [7]. (Green shade: Met the criteria for 'fully replicable'. Red shade: Met the criteria for not 'fully replicable'. Black shade: Met the criteria for not 'fully replicable' and 'meaningfully' different.)

| | Intervention | | Control | | | | | | |
|---|---|---|---|---|---|---|---|---|---|
| | Events | Total | Events | Total | OR | 95% CI | Screening discrepancy | Data discrepancy | Reason for discrepancy |
| Original reviewers | | | | | | | | | |
| Hung et al. (2015) | 67 | 76 | 68 | 75 | 0.77 | (0.27, 2.18) | - | - | - |
| Miehlke et al. (2006) | 54 | 73 | 50 | 72 | 1.25 | (0.61, 2.58) | - | - | - |
| Navarro-Jarabo et al. (2007) | 20 | 45 | 38 | 54 | 0.34 | (0.15, 0.77) | - | - | - |
| Perri et al. (2001) | 69 | 90 | 60 | 90 | 1.64 | (0.85, 3.17) | - | - | - |
| [Random effects model] | - | - | - | - | 0.89 | (0.44, 1.79) | | | |
| Replicators (Blinded) | | | | | | | | | |
| Hung et al. (2015) | 67 | 76 | 68 | 75 | 0.77 | (0.27, 2.18) | No | No | - |
| Miehlke et al. (2006) | 54 | 73 | 50 | 72 | 1.25 | (0.61, 2.58) | No | No | - |
| Navarro-Jarabo et al. (2007) | 20 | 45 | 38 | 54 | 0.34 | (0.15, 0.77) | No | No | - |
| Perri et al. (2001) | 69 | 90 | 30 | 45 | 1.64 | (0.75, 3.62) | No | Yes | Reviewer extraction error |
| [Random effects model (R1)] | - | - | - | - | 0.87 | (0.43, 1.76) | | | |
| [Random effects model (R2)] | - | - | - | - | 0.87 | (0.43, 1.76) | | | |
| Replicators (Unblinded) | | | | | | | | | |
| Hung et al. (2015) | 67 | 76 | 68 | 75 | 0.77 | (0.27, 2.18) | No | No | - |
| Miehlke et al. (2006) | 54 | 73 | 50 | 72 | 1.25 | (0.61, 2.58) | No | No | - |
| Navarro-Jarabo et al. (2007) | 20 | 45 | 38 | 54 | 0.34 | (0.15, 0.77) | No | No | - |
| Perri et al. (2001) | 69 | 90 | 30 | 45 | 1.64 | (0.75, 3.62) | No | Yes | Reviewer extraction error |
| [Random effects model (R1)] | - | - | - | - | 0.87 | (0.43, 1.76) | | | |
| [Random effects model (R2)] | - | - | - | - | 0.87 | (0.43, 1.76) | | | |

Supplementary Figure 5. Results of the original and replicated index meta-analyses for the review by Goldberg et al. (2020) [8]. (Green shade: Met the criteria for 'fully replicable'. Red shade: Met the criteria for not 'fully replicable'. Black shade: Met the criteria for not 'fully replicable' and 'meaningfully' different.)

| | Intervention | | | Control | | | | | | | |
|---|---|---|---|---|---|---|---|---|---|---|---|
| | Mean | SD | Total | Mean | SD | Total | Cohen's d | 95% CI | Screening discrepancy | Data discrepancy | Reason for discrepancy |
| Original reviewers | | | | | | | | | | | |
| Gasser et al. (2014) | - | - | - | - | - | - | 2.22 | (0.91, 3.53)* | - | - | - |
| Griffiths et al. (2016) | - | - | - | - | - | - | 0.86 | (0.24, 1.48)* | - | - | - |
| Grob et al. (2011) | - | - | - | - | - | - | 0.94 | (-0.06, 1.94)* | - | - | - |
| Palhano-Fontes et al. (2019) | - | - | - | - | - | - | 1.20 | (0.52, 1.88)* | - | - | - |
| Ross et al. (2016) | - | - | - | - | - | - | 0.98 | (0.27, 1.69)* | - | - | - |
| [Random effects model] | - | - | - | - | - | - | 1.08 | (0.74, 1.43) | | | |
| Replicators (Blinded) | | | | | | | | | | | |
| Gasser et al. (2014) | 6.6 | 37.7 | 8 | 3.4 | 24.3 | 3 | 0.09 | (-1.24, 1.42) | No | Yes | Incomplete data selection methods |
| Griffiths et al. (2016) | 11.6 | 5.8 | 26 | 8.5 | 5.4 | 25 | 0.55 | (-0.01, 1.11) | No | Yes | Incomplete data selection methods |
| Grob et al. (2011) | 9.7 | 7.8 | 6 | 0.1 | 8.4 | 6 | 1.21 | (-0.04, 2.46) | No | Yes | Incomplete data selection methods |
| Palhano-Fontes et al. (2019) | 14.4 | 6.6 | 14 | 2.8 | 6.4 | 15 | 1.77 | (0.90, 2.64) | No | Yes | Incomplete data selection methods |
| Ross et al. (2016) | 11.7 | 10.3 | 14 | 5.5 | 10.1 | 15 | 0.61 | (-0.14, 1.35) | No | Yes | Incomplete data selection methods |
| [Random effects model (R1)] | - | - | - | - | - | - | 0.84 | (0.31, 1.37) | | | |
| [Random effects model (R2)] | - | - | - | - | - | - | 0.84 | (0.31, 1.38) | | | |
| Replicators (Unblinded) | | | | | | | | | | | |
| Gasser et al. (2014) | 6.6 | 37.7 | 8 | 3.4 | 24.3 | 3 | 0.09 | (-1.24, 1.42) | No | Yes | Incomplete data selection methods |
| Griffiths et al. (2016) | 11.6 | 5.8 | 26 | 8.5 | 5.4 | 25 | 0.55 | (-0.01, 1.11) | No | Yes | Incomplete data selection methods |
| Grob et al. (2011) | 9.7 | 7.8 | 6 | 0.1 | 8.4 | 6 | 1.21 | (-0.04, 2.46) | No | Yes | Incomplete data selection methods |
| Palhano-Fontes et al. (2019) | 14.4 | 6.6 | 14 | 2.8 | 6.4 | 15 | 1.77 | (0.90, 2.64) | No | Yes | Incomplete data selection methods |
| Ross et al. (2016) | 11.7 | 10.3 | 14 | 5.5 | 10.1 | 15 | 0.61 | (-0.14, 1.35) | No | Yes | Incomplete data selection methods |
| [Random effects model (R1)] | - | - | - | - | - | - | 0.84 | (0.31, 1.37) | | | |
| [Random effects model (R2)] | - | - | - | - | - | - | 0.84 | (0.31, 1.38) | | | |

Supplementary Figure 6. Results of the original and replicated index meta-analyses for the review by Ioannou et al. (2021) [9]. (Green shade: Met the criteria for 'fully replicable'. Red shade: Met the criteria for not 'fully replicable'. Black shade: Met the criteria for not 'fully replicable' and 'meaningfully' different.)

| | Intervention | | | Control | | | | | Screening discrepancy | Data discrepancy | Reason for discrepancy |
|---|---|---|---|---|---|---|---|---|---|---|---|
| | Mean | SD | Total | Mean | SD | Total | Hedge's g | 95% CI | | | |
| Original reviewers | | | | | | | | | | | |
| Benedetti et al. (1997) | 9.6 | 11.4 | 5 | 16.4 | 12.9 | 5 | -0.51 | (-1.78, 0.77) | - | - | - |
| Elsenga et al. (1982) | 21.5 | 1.6 | 10 | 23.2 | 1.6 | 10 | -1.03 | (-1.98, -0.09) | - | - | - |
| Kragh et al. (2017a) | 17.4 | 5.2 | 32 | 20.2 | 5.5 | 32 | -0.52 | (-1.02, -0.02) | - | - | - |
| Kundermann et al. (2008) | 16.3 | 6.6 | 9 | 17.8 | 9.4 | 10 | -0.17 | (-1.07, 0.73) | - | - | - |
| Reynolds et al. (2005) | 15.1 | 3.9 | 27 | 12.1 | 4.1 | 26 | 0.74 | (0.18, 1.29) | - | - | - |
| Wu et al. (2009) | 10.2 | 7.3 | 32 | 14.4 | 8.0 | 17 | -0.55 | (-1.15, 0.05) | - | - | - |
| [Random effects model] | - | - | - | - | - | - | -0.29 | (-0.84, 0.25) | | | |
| Replicators (Blinded) | | | | | | | | | | | |
| Benedetti et al. (1997) | 14.1 | 10.7 | 5 | 22.1 | 12.7 | 5 | -0.61 | (-1.90, 0.67) | No | Yes | Reviewer extraction error |
| Elsenga et al. (1982) | NI | NI | NI | NI | NI | NI | NI | NI | Yes | - | Incomplete eligibility criteria |
| Kragh et al. (2017a) | NI | NI | NI | NI | NI | NI | NI | NI | Yes | - | Unclear eligibility criteria |
| Kundermann et al. (2008) | 16.3 | 6.6 | 9 | 17.8 | 9.5 | 10 | -0.17 | (-1.08, 0.73) | No | Yes | Data extraction from plot |
| Reynolds et al. (2005) | NI | NI | NI | NI | NI | NI | NI | NI | Yes | - | Unable to obtain data |
| Wu et al. (2009) | NI | NI | NI | NI | NI | NI | NI | NI | Yes | - | Incomplete eligibility criteria |
| [Random effects model (R1)] | - | - | - | - | - | - | -0.32 | (-1.06, 0.42) | | | |
| [Random effects model (R2)] | - | - | - | - | - | - | -0.33 | (-1.02, 0.36) | | | |
| Replicators (Unblinded) | | | | | | | | | | | |
| Benedetti et al. (1997) | 14.1 | 10.7 | 5 | 22.1 | 12.7 | 5 | -0.61 | (-1.90, 0.67) | No | Yes | Reviewer extraction error |
| Elsenga et al. (1982) | 21.5 | 1.6 | 10 | 23.2 | 1.6 | 10 | -1.03 | (-1.98, -0.09) | No | No | - |
| Kragh et al. (2017a) | NI | NI | NI | NI | NI | NI | NI | NI | Yes | - | Unclear eligibility criteria |
| Kundermann et al. (2008) | 16.3 | 6.6 | 9 | 17.8 | 9.5 | 10 | -0.17 | (-1.08, 0.73) | No | Yes | Data extraction from plot |
| Reynolds et al. (2005) | 15.1 | 3.9 | 27 | 12.1 | 4.1 | 26 | 0.74 | (0.18, 1.29) | No | No | - |
| Wu et al. (2009) | 10.2 | 7.3 | 32 | 14.4 | 8.0 | 17 | -0.55 | (-1.15, 0.05) | No | No | - |
| [Random effects model (R1)] | - | - | - | - | - | - | -0.27 | (-0.96, 0.43) | | | |
| [Random effects model (R2)] | - | - | - | - | - | - | -0.28 | (-0.96, 0.41) | | | |

Supplementary Figure 7. Results of the original and replicated index meta-analyses for the review by Jakubczuk et al. (2020) [10]. (Green shade: Met the criteria for 'fully replicable'. Red shade: Met the criteria for not 'fully replicable'. Black shade: Met the criteria for not 'fully replicable' and 'meaningfully' different.)

| | Intervention | | | Control | | | | | | | |
|---|---|---|---|---|---|---|---|---|---|---|---|
| | Mean | SD | Total | Mean | SD | Total | Cohen's d | 95% CI | Screening discrepancy | Data discrepancy | Reason for discrepancy |
| Original reviewers | | | | | | | | | | | |
| Alizadeh et al. (2017) | - | - | - | - | - | - | 8.24 | (6.63, 9.86) | - | - | - |
| Nasseri et al. (2017) | - | - | - | - | - | - | 0.14 | (-0.36, 0.65) | - | - | - |
| Saraf-Bank et al. (2019) | - | - | - | - | - | - | 0.28 | (-0.23, 0.79) | - | - | - |
| [Random effects model] | - | - | - | - | - | - | 2.70 | (0.06, 5.34) | | | |
| Replicators (Blinded) | | | | | | | | | | | |
| Gasser et al. (2014) | 2.0 | 0.1 | 28 | 1.4 | 0.1 | 28 | 8.24 | (6.63, 9.86) | No | No | - |
| Griffiths et al. (2016) | 208.0 | 91.7 | 31 | 196.1 | 72.6 | 30 | 0.14 | (-0.36, 0.65) | No | No | - |
| Grob et al. (2011) | 230.0 | 40.0 | 30 | 220.0 | 30.0 | 30 | 0.28 | (-0.23, 0.79) | No | No | - |
| [Random effects model (R1)] | - | - | - | - | - | - | 2.68 | (0.08, 5.28) | | | |
| [Random effects model (R2)] | - | - | - | - | - | - | 2.70 | (0.06, 5.34) | | | |
| Replicators (Unblinded) | | | | | | | | | | | |
| Gasser et al. (2014) | 2.0 | 0.1 | 28 | 1.4 | 0.1 | 28 | 8.24 | (6.63, 9.86) | No | No | - |
| Griffiths et al. (2016) | 208.0 | 91.7 | 31 | 196.1 | 72.6 | 30 | 0.14 | (-0.36, 0.65) | No | No | - |
| Grob et al. (2011) | 230.0 | 40.0 | 30 | 220.0 | 30.0 | 30 | 0.28 | (-0.23, 0.79) | No | No | - |
| [Random effects model (R1)] | - | - | - | - | - | - | 2.68 | (0.08, 5.28) | | | |
| [Random effects model (R2)] | - | - | - | - | - | - | 2.70 | (0.06, 5.34) | | | |

Supplementary Figure 8. Results of the original and replicated index meta-analyses for the review by Minozzi et al. (2020) [11]. (Green shade: Met the criteria for 'fully replicable'. Red shade: Met the criteria for not 'fully replicable'. Black shade: Met the criteria for not 'fully replicable' and 'meaningfully' different.)

| | Intervention | | Control | | | | Screening discrepancy | Data discrepancy | Reason for discrepancy |
|---|---|---|---|---|---|---|---|---|---|
| | Events | Total | Events | Total | RR | 95% CI | | | |
| Original reviewers | | | | | | | | | |
| Fischer et al. (2006) | 3 | 9 | 1 | 9 | 3.00 | (0.38, 23.68) | - | - | - |
| Jones et al. (2005) | 4 | 15 | 6 | 15 | 0.67 | (0.23, 1.89) | - | - | - |
| Jones et al. (2010) | 16 | 89 | 28 | 86 | 0.55 | (0.32, 0.95) | - | - | - |
| [Random effects model] | - | - | - | - | 0.66 | (0.37, 1.20) | | | |
| Replicators (Blinded) | | | | | | | | | |
| Fischer et al. (2006) | 3 | 9 | 1 | 9 | 3.00 | (0.38, 23.68) | No | No | - |
| Jones et al. (2005) | 4 | 15 | 6 | 15 | 0.67 | (0.23, 1.89) | No | No | - |
| Jones et al. (2010) | 16 | 89 | 28 | 86 | 0.55 | (0.32, 0.95) | No | No | - |
| [Random effects model (R1)] | - | - | - | - | 0.66 | (0.37, 1.20) | | | |
| [Random effects model (R2)] | - | - | - | - | 0.66 | (0.37, 1.19) | | | |
| Replicators (Unblinded) | | | | | | | | | |
| Fischer et al. (2006) | 3 | 9 | 1 | 9 | 3.00 | (0.38, 23.68) | No | No | - |
| Jones et al. (2005) | 4 | 15 | 6 | 15 | 0.67 | (0.23, 1.89) | No | No | - |
| Jones et al. (2010) | 16 | 89 | 28 | 86 | 0.55 | (0.32, 0.95) | No | No | - |
| [Random effects model (R1)] | - | - | - | - | 0.66 | (0.37, 1.20) | | | |
| [Random effects model (R2)] | - | - | - | - | 0.66 | (0.37, 1.19) | | | |

Supplementary Figure 9. Results of the original and replicated index meta-analyses for the review by Wang et al. (2020) [12]. (Green shade: Met the criteria for 'fully replicable'. Red shade: Met the criteria for not 'fully replicable'. Black shade: Met the criteria for not 'fully replicable' and 'meaningfully' different.)

| | HR | 95% CI | Screening discrepancy | Data discrepancy | Reason for discrepancy |
|---|---|---|---|---|---|
| Original reviewers | | | | | |
| Akiyama et al. (1994) | NI | NI | - | - | - |
| Fan et al. (2019) | 1.02 | (0.66, 1.57) | - | - | - |
| Fujita et al. (1995) | 1.13 | (0.67, 1.92) | - | - | - |
| Fujita et al. (2003) | NI | NI | - | - | - |
| Igaki et al. (2004) | 1.29 | (0.77, 2.15) | - | - | - |
| Koterazawa et al. (2019) | NI | NI | - | - | - |
| Li et al. (2012) | 0.88 | (0.63, 1.23) | - | - | - |
| Shao et al. (2018) | 0.94 | (0.71, 1.24) | - | - | - |
| Shim et al. (2010) | 1.35 | (0.71, 2.58) | - | - | - |
| Tabira et al. (1999) | 1.20 | (0.65, 2.20) | - | - | - |
| Zhang et al. (2008) | 1.31 | (0.82, 2.10) | - | - | - |
| [Fixed effect model] | 1.05 | (0.90, 1.21) | | | |
| Replicators (Blinded) | | | | | |
| Akiyama et al. (1994) | 1.54 | (1.22, 1.95) | Yes | - | Coordinator extraction error |
| Fan et al. (2019) | 1.01 | (0.63, 1.62) | No | Trivial | Software-related |
| Fujita et al. (1995) | 1.19 | (0.77, 1.83) | No | Yes | Data extraction from plot |
| Fujita et al. (2003) | 0.36 | (0.25, 0.53) | Yes | - | Coordinator extraction error |
| Igaki et al. (2004) | 1.25 | (0.80, 1.96) | No | Yes | Data extraction from plot |
| Koterazawa et al. (2019) | 1.15 | (0.68, 1.95) | Yes | - | Coordinator extraction error |
| Li et al. (2012) | 0.88 | (0.62, 1.26) | No | Yes | Data extraction from plot |
| Shao et al. (2018) | 0.93 | (0.72, 1.19) | No | Trivial | Software-related |
| Shim et al. (2010) | 1.15 | (0.64, 2.07) | No | Yes | Data extraction from plot |
| Tabira et al. (1999) | 1.52 | (0.89, 2.57) | No | Yes | Data extraction from plot |
| Zhang et al. (2008) | 1.30 | (0.86, 1.96) | No | Yes | Data extraction from plot |
| [Fixed effect model (R1)] | 1.06 | (0.94, 1.18) | | | |
| [Fixed effect model (R2)] | 1.06 | (0.95, 1.18) | | | |
| Replicators (Unblinded) | | | | | |
| Akiyama et al. (1994) | NI | NI | No | - | - |
| Fan et al. (2019) | 1.01 | (0.63, 1.62) | No | Trivial | Software-related |
| Fujita et al. (1995) | 1.19 | (0.77, 1.83) | No | Yes | Data extraction from plot |
| Fujita et al. (2003) | NI | NI | No | - | - |
| Igaki et al. (2004) | 1.25 | (0.80, 1.96) | No | Yes | Data extraction from plot |
| Koterazawa et al. (2019) | NI | NI | No | - | - |
| Li et al. (2012) | 0.88 | (0.62, 1.26) | No | Yes | Data extraction from plot |
| Shao et al. (2018) | 0.93 | (0.72, 1.19) | No | Trivial | Software-related |
| Shim et al. (2010) | 1.15 | (0.64, 2.07) | No | Yes | Data extraction from plot |
| Tabira et al. (1999) | 1.52 | (0.89, 2.57) | No | Yes | Data extraction from plot |
| Zhang et al. (2008) | 1.30 | (0.86, 1.96) | No | Yes | Data extraction from plot |
| [Fixed effect model (R1)] | 1.07 | (0.93, 1.23) | | | |
| [Fixed effect model (R2)] | 1.07 | (0.93, 1.23) | | | |

Supplementary Figure 10. Results of the original and replicated index meta-analyses for the review by Yekeduz et al. (2020) [13]. (Green shade: Met the criteria for 'fully replicable'. Red shade: Met the criteria for not 'fully replicable'. Black shade: Met the criteria for not 'fully replicable' and 'meaningfully' different.)

| | aOR | 95% CI | Screening discrepancy | Data discrepancy | Reason for discrepancy |
|---|---|---|---|---|---|
| Original reviewers | | | | | |
| Kuderer et al. (2000) | 1.47 | (0.84, 2.57) | - | - | - |
| Lee et al. (2020a) | 2.09 | (1.09, 4.01) | - | - | - |
| Lee et al. (2020b) | NI | NI | - | - | - |
| Yang et al. (2020) | 3.51 | (1.16, 10.62) | - | - | - |
| Yarza et al. (2020) | 1.60 | (0.40, 6.40) | - | - | - |
| [Random effects model] | 1.85 | (1.26, 2.71) | | | |
| Replicators (Blinded) | | | | | |
| Kuderer et al. (2000) | 1.47 | (0.84, 2.56) | No | Trivial | Software-related |
| Lee et al. (2020a) | 2.09 | (1.09, 4.08) | No | Trivial | Software-related |
| Lee et al. (2020b) | 1.18 | (0.81, 1.72) | Yes | - | Replicator screening error |
| Yang et al. (2020) | 3.51 | (1.16, 10.59) | No | Trivial | Software-related |
| Yarza et al. (2020) | 1.60 | (0.40, 6.33) | No | Trivial | Software-related |
| [Random effects model (R1)] | 1.53 | (1.12, 2.10) | | | |
| [Random effects model (R2)] | 1.53 | (1.12, 2.10) | | | |
| Replicators (Unblinded) | | | | | |
| Kuderer et al. (2000) | 1.47 | (0.84, 2.56) | No | Trivial | Software-related |
| Lee et al. (2020a) | 2.09 | (1.09, 4.08) | No | Trivial | Software-related |
| Lee et al. (2020b) | NI | NI | No | - | - |
| Yang et al. (2020) | 3.51 | (1.16, 10.59) | No | Trivial | Software-related |
| Yarza et al. (2020) | 1.60 | (0.40, 6.33) | No | Trivial | Software-related |
| [Random effects model (R1)] | 1.85 | (1.26, 2.71) | | | |
| [Random effects model (R2)] | 1.85 | (1.26, 2.71) | | | |

Supplementary Figure 11. Results of the original and replicated index meta-analyses for the review by Zhou et al. (2021) [14]. (Green shade: Met the criteria for 'fully replicable'. Red shade: Met the criteria for not 'fully replicable'. Black shade: Met the criteria for not 'fully replicable' and 'meaningfully' different.) *Confidence intervals from the original review were estimated in R.

| | Intervention | | | Control | | | | | Screening discrepancy | Data discrepancy | Reason for discrepancy |
|---|---|---|---|---|---|---|---|---|---|---|---|
| | Mean | SD | Total | Mean | SD | Total | Hedge's g | 95% CI | | | |
| Original reviewers | | | | | | | | | | | |
| Hoseini et al. (2016) | NI | NI | NI | NI | NI | NI | NI | NI | - | - | - |
| Kajagar et al. (2012) | - | - | 34 | - | - | 34 | 5.21 | (4.22, 6.20)* | - | - | - |
| Kaviani et al. (2011) | - | - | 13 | - | - | 10 | 2.00 | (1.02, 2.98)* | - | - | - |
| Mathur et al. (2017) | - | - | 15 | - | - | 15 | 2.98 | (1.95, 4.01)* | - | - | - |
| Minatel et al. (2009) | - | - | 7 | - | - | 7 | 2.29 | (0.99, 3.59)* | - | - | - |
| Naidu et al. (2005) | - | - | 8 | - | - | 8 | 4.17 | (2.45, 5.89)* | - | - | - |
| Zhang et al. (2013) | - | - | 42 | - | - | 42 | 0.45 | (0.02, 0.88)* | - | - | - |
| [Random effects model] | - | - | - | - | - | - | 2.81 | (1.14, 4.48)* | | | |
| Replicators (Blinded) | | | | | | | | | | | |
| Hoseini et al. (2016) | 44.0 | 6.4 | 15 | 6.5 | 8.0 | 12 | 5.11 | (3.46, 6.76) | Yes | - | Unclear |
| Kajagar et al. (2012) | 40.2 | 6.3 | 34 | 11.9 | 4.3 | 34 | 5.21 | (4.19, 6.23) | No | Trivial | Software-related |
| Kaviani et al. (2011) | 47.5 | 9.0 | 13 | 29.4 | 7.6 | 10 | 2.07 | (1.02, 3.12) | No | Trivial | Software-related |
| Mathur et al. (2017) | 37.3 | 8.8 | 15 | 15.2 | 5.4 | 15 | 2.94 | (1.87, 4.01) | No | Trivial | Software-related |
| Minatel et al. (2009) | NI | NI | NI | NI | NI | NI | NI | NI | Yes | - | Ambiguous eligiblity criteria |
| Naidu et al. (2005) | 41.3 | 15.7 | 8 | 4.6 | 6.0 | 8 | 2.92 | (1.40, 4.43) | No | Yes | Unclear |
| Zhang et al. (2013) | NI | NI | NI | NI | NI | NI | NI | NI | Yes | - | Unclear eligiblity criteria |
| [Random effects model (R1)] | | | | | | | 3.62 | (2.31, 4.93) | | | |
| [Random effects model (R2)] | | | | | | | 3.62 | (2.33, 4.91) | | | |
| Replicators (Unblinded) | | | | | | | | | | | |
| Hoseini et al. (2016) | 44.0 | 6.4 | 15 | 6.5 | 8.0 | 12 | 5.11 | (3.46, 6.76) | Yes | - | Unclear |
| Kajagar et al. (2012) | 40.2 | 6.3 | 34 | 11.9 | 4.3 | 34 | 5.21 | (4.19, 6.23) | No | Trivial | Software-related |
| Kaviani et al. (2011) | 47.5 | 9.0 | 13 | 29.4 | 7.6 | 10 | 2.07 | (1.02, 3.12) | No | Trivial | Software-related |
| Mathur et al. (2017) | 37.3 | 8.8 | 15 | 15.2 | 5.4 | 15 | 2.94 | (1.87, 4.01) | No | Trivial | Software-related |
| Minatel et al. (2009) | 44.1 | 21.6 | 7 | -8.9 | 24.7 | 7 | 2.14 | (0.74, 3.53) | No | Trivial | Software-related |
| Naidu et al. (2005) | 41.3 | 15.7 | 8 | 4.6 | 6.0 | 8 | 2.92 | (1.40, 4.43) | No | Yes | Unclear |
| Zhang et al. (2013) | NI | NI | NI | NI | NI | NI | NI | NI | Yes | - | Unclear eligiblity criteria |
| [Random effects model (R1)] | | | | | | | 3.38 | (2.21, 4.55) | | | |
| [Random effects model (R2)] | | | | | | | 3.38 | (2.22, 4.53) | | | |

Supplementary Figure 12. Results of the original and sensitivity meta-analyses for the review by AlAnouti et al. (2020) [5]. (Green shade: Met the criteria for 'fully replicable'. Red shade: Met the criteria for not 'fully replicable'. Black shade: Met the criteria for not 'fully replicable' and 'meaningfully' different.)

| | Intervention | | | Control | | | | | Screening discrepancy | Data discrepancy | Reason for discrepancy |
|---|---|---|---|---|---|---|---|---|---|---|---|
| | Mean | SD | Total | Mean | SD | Total | MD | 95% CI | | | |
| Original reviewers | | | | | | | | | | | |
| Farag et al. (2019) | 233.8 | 97.0 | 24 | 158.6 | 35.4 | 25 | 75.20 | (33.99, 116.41) | - | - | - |
| Makariou et al. (2017) | NA | NA | NA | NA | NA | NA | NA | NA | - | - | - |
| Wongwiwatthananukit (2013) | 137.8 | 53.5 | 28 | 135.8 | 71.4 | 28 | 2.04 | (-31.00, 35.08) | - | - | - |
| Yin et al. (2016) | NI | NI | NI | NI | NI | NI | NI | NI | - | - | - |
| [Fixed effect model] | - | - | - | - | - | - | 30.67 | (4.89, 56.45) | | | |
| Replicators (Blinded) | | | | | | | | | | | |
| Farag et al. (2019) | 233.8 | 97.0 | 24 | 158.6 | 35.4 | 25 | 75.20 | (33.99, 116.41) | No | No | - |
| Makariou et al. (2017) | 139.3 | 52.9 | 25 | 143.7 | 53.3 | 25 | -4.40 | (-33.84, 25.04) | NA | NA | NA |
| Wongwiwatthananukit (2013) | 137.8 | 53.5 | 30 | 135.8 | 71.4 | 30 | 2.04 | (-29.88, 33.96) | No | Yes | Reviewer extraction error |
| Yin et al. (2016) | 250.4 | 36.3 | 61 | 254.9 | 19.5 | 62 | -4.43 | (-14.74, 5.88) | Yes | - | Coordinator extraction error |
| [Fixed effect model (R1)] | - | - | - | - | - | - | -0.09 | (-9.17, 9.00) | | | |
| Replicators (Unblinded) | | | | | | | | | | | |
| Farag et al. (2019) | 233.8 | 97.0 | 24 | 158.6 | 35.4 | 25 | 75.20 | (33.99, 116.41) | No | No | - |
| Makariou et al. (2017) | 139.3 | 52.9 | 25 | 143.7 | 53.3 | 25 | -4.40 | (-33.84, 25.04) | NA | NA | NA |
| Wongwiwatthananukit (2013) | 137.8 | 53.5 | 30 | 135.8 | 71.4 | 30 | 2.04 | (-29.88, 33.96) | No | Yes | Reviewer extraction error |
| Yin et al. (2016) | NI | NI | NI | NI | NI | NI | NI | NI | No | - | - |
| [Fixed effect model (R1)] | - | - | - | - | - | - | 15.12 | (-4.04, 34.28) | | | |

Supplementary Figure 13. Results of the original and sensitivity meta-analyses for the review by Wang et al. (2020) [12]. (Green shade: Met the criteria for 'fully replicable'. Red shade: Met the criteria for not 'fully replicable'. Black shade: Met the criteria for not 'fully replicable' and 'meaningfully' different.)

| | HR | 95% CI | Screening discrepancy | Data discrepancy | Reason for discrepancy |
|---|---|---|---|---|---|
| Original reviewers | | | | | |
| Akiyama et al. (1994) | NI | NI | - | - | - |
| Fan et al. (2019) | 1.02 | (0.66, 1.57) | - | - | - |
| Fujita et al. (1995) | 1.13 | (0.67, 1.92) | - | - | - |
| Fujita et al. (2003) | NI | NI | - | - | - |
| Igaki et al. (2004) | 1.29 | (0.77, 2.15) | - | - | - |
| Koterazawa et al. (2019) | NI | NI | - | - | - |
| Li et al. (2012) | 0.88 | (0.63, 1.23) | - | - | - |
| Shao et al. (2018) | 0.94 | (0.71, 1.24) | - | - | - |
| Shim et al. (2010) | 1.35 | (0.71, 2.58) | - | - | - |
| Tabira et al. (1999) | 1.20 | (0.65, 2.20) | - | - | - |
| Zhang et al. (2008) | 1.31 | (0.82, 2.10) | - | - | - |
| [Fixed effect model] | 1.05 | (0.90, 1.21) | | | |
| Replicators (Blinded) | | | | | |
| Akiyama et al. (1994) | Exc | Exc | NA | NA | NA |
| Fan et al. (2019) | 1.01 | (0.63, 1.62) | No | Trivial | Software-related |
| Fujita et al. (1995) | Exc | Exc | NA | NA | NA |
| Fujita et al. (2003) | 0.36 | (0.25, 0.53) | Yes | - | Coordinator extraction error |
| Igaki et al. (2004) | Exc | Exc | NA | NA | NA |
| Koterazawa et al. (2019) | 1.15 | (0.68, 1.95) | Yes | - | Coordinator extraction error |
| Li et al. (2012) | 0.88 | (0.62, 1.26) | No | Yes | Data extraction from plot |
| Shao et al. (2018) | 0.93 | (0.72, 1.19) | No | Trivial | Software-related |
| Shim et al. (2010) | 1.15 | (0.64, 2.07) | No | Yes | Data extraction from plot |
| Tabira et al. (1999) | 1.52 | (0.89, 2.57) | No | Yes | Data extraction from plot |
| Zhang et al. (2008) | Exc | Exc | NA | NA | NA |
| [Fixed effect model (R1)] | 1.01 | (0.86, 1.19) | | | |
| Replicators (Unblinded) | | | | | |
| Akiyama et al. (1994) | NI | NI | No | - | - |
| Fan et al. (2019) | 1.01 | (0.63, 1.62) | No | Trivial | Software-related |
| Fujita et al. (1995) | Exc | Exc | NA | NA | NA |
| Fujita et al. (2003) | NI | NI | No | - | - |
| Igaki et al. (2004) | Exc | Exc | NA | NA | NA |
| Koterazawa et al. (2019) | NI | NI | No | - | - |
| Li et al. (2012) | 0.88 | (0.62, 1.26) | No | Yes | Data extraction from plot |
| Shao et al. (2018) | 0.93 | (0.72, 1.19) | No | Trivial | Software-related |
| Shim et al. (2010) | 1.15 | (0.64, 2.07) | No | Yes | Data extraction from plot |
| Tabira et al. (1999) | 1.52 | (0.89, 2.57) | No | Yes | Data extraction from plot |
| Zhang et al. (2008) | Exc | Exc | NA | NA | NA |
| [Fixed effect model (R1)] | 0.99 | (0.84, 1.18) | | | |
